# Unraveling Genetic Variants underlying Schizophrenia Phenotypes: An Original GWAS in Hong Kong Chinese with Cross-Ethnic Meta-Analysis and Predictive Modeling

**DOI:** 10.64898/2025.12.24.25342953

**Authors:** ST Rao, Kenneth CY Wong, Shuai Zhi, Zoe ZY Zheng, Perry BM Leung, Benedict KW Lee, Eric FC Cheung, Raymond CK Chan, Karen KY Ho, Karen SY Hung, Tomy CK Hui, Tao Li, Pak C Sham, Simon SY Lui, Hon-Cheong So

**Author notes:** These authors contributed equally to this work (ST Rao, Kenneth CY Wong).

## Abstract

**Background:** Schizophrenia (SCZ) is a highly heritable and heterogeneous disorder with diverse clinical presentations and cognitive deficits. The specific genetic variants contributing to this variability remain largely unknown. This study aims to uncover the genetic bases of various clinical phenotypes such as age at onset (AAO), positive/negative symptoms, self-harm and aggression in SCZ using genome-wide association studies (GWAS). Few large-scale GWAS have explored these phenotypes, especially in non-Europeans. Understanding the genetics may reveal new therapeutic targets in the future. Additionally, it is unclear whether the genetics of SCZ phenotypes overlap with the genetic risk for SCZ itself or other psychiatric disorders. Previous studies using polygenic risk scores have provided limited insights.

**Aims and Objectives:** This study aims to: (i) identify genetic variants associated with SCZ clinical phenotypes using GWAS, (ii) decipher the genetic relationship between SCZ phenotypes and other psychiatric disorders via PRS, (iii) gain biological insights into SCZ phenotypes through gene and pathway analyses of GWAS data, and (iv) predict the severity of SCZ phenotypes using a machine learning model that combines clinical and genomic factors.

**Methods:** This observational study is primarily based on two cohorts of SCZ patients from Hong Kong, with genotyping data collected. A third cohort from dbGaP was used to study genetics of AAO. Clinical phenotypes such as self-harm, aggression, hospitalization, and AAO were collected. PRS were calculated for psychiatric and cognitive traits. GWAS were conducted for the phenotypes within each cohort and meta-analysed. Gene-based analyses identified associated genes, which were then subjected to pathway enrichment analysis. Machine learning models were built using PRS and clinical variables to predict relevant outcomes such as hospitalization risk.

**Results:** GWAS identified several significant genetic variants associated with SCZ phenotypes across the three cohorts. A variant rs60648049 was associated with hospitalization count in cohort 1. In cohort 2, a variant was linked to age at onset (AAO). The dbGaP cohort revealed two variants significantly associated with AAO. Meta-analysis across cohorts identified three variants ( rs144645158, rs142498233 and rs185213255)) significantly associated with AAO. Gene-based and transcriptome-wide analyses implicated genes in pathways related to synaptic function, neurodevelopment, and immune processes. Males, lower education, and genetic factors like lower IQ PRS were associated with increased aggressive behaviours. Self-harm was linked to lower education and ADHD PRS. Earlier AAO correlated with male sex, higher education, and PRS for bipolar disorder and major depression. Machine learning models predicted hospitalization status with an area under the receiver operator characteristic curve of 63.4%, with AAO and aggressive behaviour as top predictors. Models were also built to predict the number of hospitalizations (RMSE=1.067) and total hospitalization duration (RMSE=3.063 months), with AAO, self-harm and aggressive behaviour being significant predictors of greater severity.

**Conclusions and Implications:** This GWAS examining clinical profiles in Chinese SCZ patients, combined with European samples, identified several genetic loci associated with age at onset. Nominal trends indicating genetic overlaps with psychiatric disorders, related traits, and cognitive functions suggest shared pathophysiology and potential therapeutic approaches. Machine learning models showed moderate accuracy in predicting SCZ severity. Collectively, these findings enhance our understanding of SCZ’s genetic architecture, and provide a foundation for future research and clinical strategies aimed at improving patient outcomes.

## Introduction

Schizophrenia (SCZ) is a highly heritable illness, ranking among the top leading causes of disability worldwide [1]. Genome-wide association studies (GWAS) have been instrumental in unravelling the genetics of SCZ. However, our understanding of SCZ is far from perfect. Importantly, SCZ has long been recognized as a highly heterogeneous condition; patients may differ substantially in their clinical presentations and prognosis. As no reliable biomarkers are available, the diagnosis of SCZ is unlikely to reflect a single biological entity with uniform etiology. Genetic factors have been suggested as an important contributor to variations among SCZ patients. Family studies showed patients with a family history of SCZ had higher levels of negative symptoms [2]. Affected sibling pairs had also been shown to be correlated in clinical features and age at onset (AAO) [2].

Here we aim to study the genetic bases of a variety of clinical phenotypes in SCZ using GWAS. For clinical traits, we proposed to study age at onset (AAO), positive/negative symptoms and history of deliberate self-harm and aggressive acts. For cognitive phenotypes, we included domains including intelligence, memory, attention and executive functions. Although antipsychotics are generally effective for positive symptoms, their benefits in other domains, including negative symptoms, are limited or questionable [3]. Deciphering the genetic basis of these symptoms may lead to the discovery of new therapeutic targets and interventions. Based on the GWAS results and clinical risk factors, we developed a machine-learning prediction model to assess the severity of phenotypes of an SCZ patient.

### Genomic studies on clinical phenotypes

Candidate gene studies on clinical features of SCZ were reviewed by Ritsner [4]. However, few findings have strong evidence. GWAS provides a systematic and unbiased approach to uncovering new disease genes. However, relatively few genome-wide studies have been conducted for clinical phenotypes of SCZ patients. Fanous et al. [2] conducted the first GWAS on three symptom factor scores (positive, negative/disorganized, mood) in SCZ [5]. A GWAS of derived positive/negative symptoms was also performed by Edwards et al. [6]. While no single nucleotide polymorphisms (SNPs) achieved genome-wide significance, several genes/pathways were significantly associated with symptom measures [6]. For AAO, two previous GWAS were performed in Caucasians [7, 8]. Concerning suicidal behaviour in SCZ, Bani-Fatemi et al. [9] performed the first GWAS with a modest sample size (N=121). However, no studies on clinical phenotypes have been performed in the Chinese population. Allele frequencies and linkage disequilibrium (LD) patterns may vary between ethnic groups, rendering some causal variants more readily discovered in certain populations. Also, the varied genetic and environmental backgrounds of different populations may lead to heterogeneity in effect sizes.

### Genetic relationship of schizophrenia phenotypes with psychiatric disorders/traits

Another intriguing question to ask is: are the “disease genes” of SCZ or other disorders the same as those “symptom/phenotype genes”? A few studies have tried to address this issue by assessing whether the polygenic risk score (PRS) of SCZ predicts symptom dimensions [10]. However, these studies only use less than a hundred genome-wide significant SNPs to construct the PRS, which may not be optimal [11]. In addition, previous studies have not investigated whether other psychiatric disorders/traits (e.g., depression) have shared genetic bases with SCZ phenotypes. No studies have explored whether cognitive deficits in SCZ are genetically linked to the disorder itself or other psychiatric disorders.

### Aims and Objectives

#### To identify genetic variants associated with clinical phenotypes of SCZ

No studies on clinical phenotypes have been performed in individuals of Chinese ancestry. Allele frequencies and linkage disequilibrium (LD) patterns may vary between ethnic groups, rendering some casual variants more readily discovered in certain populations. Also, the varied genetic and environmental backgrounds of different populations may lead to heterogeneity in effect sizes. This study aimed to identify genetic variants associated with clinical phenotypes of Chinese individuals with SCZ using GWAS.

#### To decipher the genetic relationship of SCZ phenotypes with psychiatric disorders/traits

No studies have explored whether cognitive deficits in SCZ are genetically linked to the disorder itself or other psychiatric disorders. We try to address this issue by deciphering the genetic relationship between SCZ phenotypes with other psychiatric disorders/traits using PRS.

#### To gain further biological insight into schizophrenia phenotypes

We performed gene-based and pathway-based analyses on the GWAS results. Given the difficulties in finding pharmacotherapies for SCZ symptoms based on animal models alone, we believe the findings may contribute to this area.

#### To predict the severity of schizophrenia phenotypes

Accurate prediction of SCZ phenotypes is crucial for prevention and early intervention. In this study, we built a prediction model for screening patients at high risk of developing certain phenotypes, based on both clinical and genomic risk factors. We are the first to construct such a combined machine learning (ML) prediction model. We believe it will potentially be useful in delineating high-risk patients and fostering early and targeted interventions shortly.

## Methods

### Study Design

This is a naturalistic observational study comprising a mix of cross-sectional and longitudinal designs. We included patients diagnosed with SCZ or schizoaffective disorders who satisfy the selection criteria. Subjects were diagnosed before or after the study start date and they have different durations of follow-up.

### Subjects

The first two cohorts consisted of a total of 1,089 individuals of Chinese ancestry within the Hong Kong population (n = 702 in cohort 1; n = 387 in cohort 2). Patients aged 18-60 meeting DSM-IV diagnosis for SCZ were recruited. They were excluded if they had a history of substance abuse, neurological disorder, major medical illness affecting brain functioning, mental retardation, head injury with loss of consciousness or having received electroconvulsive therapy (ECT) within the past 2 years. In cohort 1, 702 subjects with clinical phenotypes were genotyped by Illumina Asian Screening Array-24 v1.0 with 659,184 probes, while 387 subjects from cohort 2 were genotyped by Illumina Human610-Quad BeadChip with 470,556 probes. Written informed consent was obtained from all participants after a comprehensive explanation of the study’s purpose and procedures. This study received approval from the Joint Chinese University of Hong Kong - New Territories East Cluster (CUHK-NTEC) Clinical Research Ethics Committee or the New Territories West Cluster Research Ethics Committee (NTW CREC).

Additionally, we included a dbGaP cohort (phs000021.v3.p2) to explore potential genetic variants associated with the AAO of SCZ. This cohort consisted of 453 SCZ patients (European-American and African-American) from United States and one from Australia, with AAO information available. Genotyping of these patients was conducted using an Affymetrix 6.0 array with 934,940 probes targeting genetic variants. All patients in this cohort were aged 18 or above at recruitment and met DSM-IV diagnosis for SCZ, and they signed written informed consent during the recruitment.

### Phenotypes

A panel of clinical parameters was collected from a review of electronic and paper case notes. The parameters included demographic data and clinical phenotypes. In cohorts 1 and 2, we aimed to assess the potential associations between the genotyped variants and several clinical phenotypes among SCZ patients. The phenotypes assessed include a history of self-harm (Yes/No), a history of aggressive acts (Yes/No), a history of violent acts (Yes/No), the categorization of SCZ episodes as either episodic or continuous, a history of hospitalization (Yes/No), the count of hospitalizations, and the AAO. Because the two hospitalization-related phenotypes serve as proxies for the severity and prognosis of SCZ, the first admission record at onset was excluded, while subsequent hospitalizations due to relapse of severe SCZ-related symptoms during the follow-up period were included. Across all three cohorts, AAO refers to the age at onset of psychotic symptoms consistent with the onset of SCZ or schizoaffective disorder. Furthermore, we examined the data distribution of the AAO across the three cohorts, as presented in **Figure 1**. In the dbGaP cohort, only the AAO phenotype was extracted for GWAS and meta-analysis.

**Figure 1.**
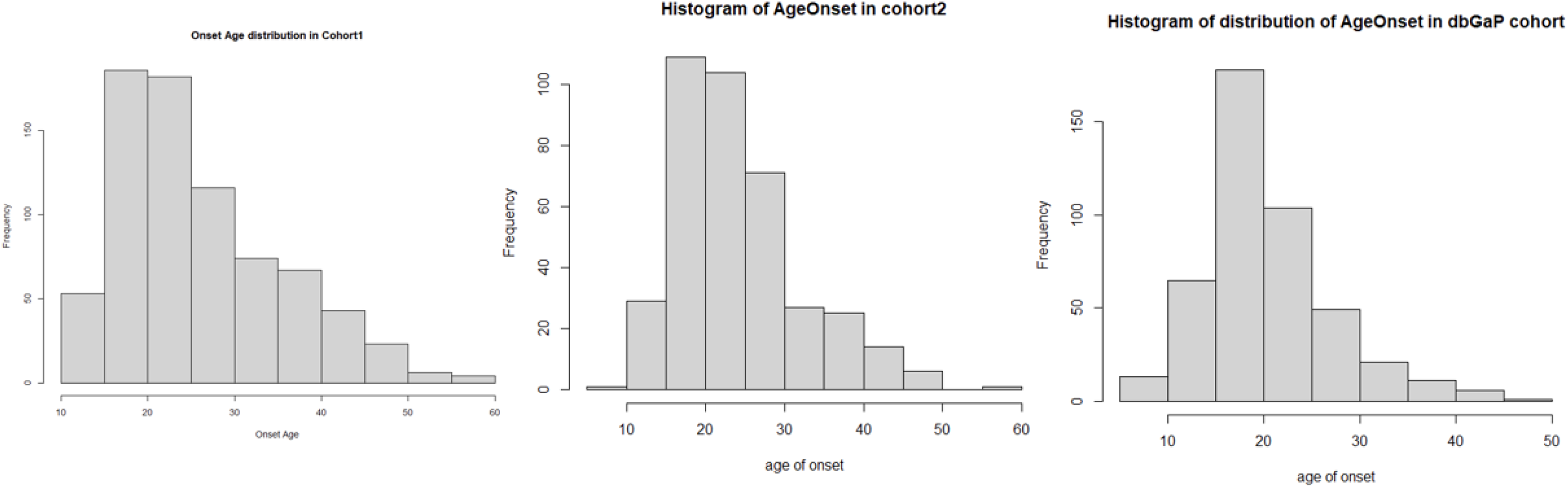
Data distribution test of the Age at onset for the three cohorts.

To study the shared genetic bases among SCZ phenotypes and the genetic overlap of these phenotypes with other psychiatric disorders, clinical phenotypes such as aggressive acts, self-harm, and AAO from cohort 1 were studied with the help of GWAS summary statistics of five psychiatric disorder - bipolar disorder (BIP), autism spectrum disorder(ASD), attention deficit hyperactivity disorder (ADHD) and major depressive disorder (MDD) downloaded from the Psychiatric Genomics Consortium (PGC). In addition, to study the genetic correlation between SCZ phenotypes and cognitive functions, GWAS summary statistics of four cognitive phenotypes - fluid intelligence score (IQ), prospective memory (PM), reaction time (RT), and memory (MEM) were downloaded from UK Biobank, as detailed in **Table 1**.

**Table 1.** Psychiatric disorders and cognitive phenotypes involved in the study of genetic overlap between these traits.

| Name | Abbr. | Database | File | Traits |
| --- | --- | --- | --- | --- |
| Bipolar disorder | BIP | PGC | NA | Psychiatric |
| Autism Spectrum disorder | ASD | PGC | NA | Psychiatric |
| Attention deficit<br>hyperactivity disorder | ADHD | PGC | NA | Psychiatric |
| Major depressive disorder | MDD | PGC | NA | Psychiatric |
| Fluid Intelligence score | IQ | UKBB | continuous-20016-both_sexes-<br>irnt.tsv.bgz | Cognitive |
| Pair matching | PM | UKBB | continuous-398-both_sexes.tsv.bgz | Cognitive |
| Reaction time | RT | UKBB | continuous-20023-both_sexes-<br>irnt.tsv.bgz | Cognitive |
| Prospective memory | MEM | UKBB | continuous-20018-both_sexes.tsv.bgz | Cognitive |

Regarding the building of a prediction model on SCZ phenotypes, a list of predictors at baseline was collected - gender, AAO, age, education level, employment, family history, immigration status, self-harm acts and aggressive acts, where gender, family history, self-harm acts and aggressive acts were treated as binary. Besides, the PRS for SCZ and other relevant psychiatric/cognitive traits including ASD, BIP, ADHD, MDD, IQ, VNR and RT were also selected as predictors in our models.

### Data cleaning and pre-processing

#### Genotyping data

Stringent quality control protocols were applied to the SNP-array genotyping data using the PLINK v1.9 program [12]. Subjects with a missing genotype rate of ≥ 10% were excluded from the analyses, while SNPs were removed if they had a missing genotype rate of ≥ 10% or a Hardy-Weinberg equilibrium (HWE) p-value < 1E−06. Subsequently, genome coordinate conversion from older genomic versions to the latest version (hg38) was performed using CrossMap [13]. The converted datasets from the two Chinese cohorts were then uploaded to the ChinaMAP Imputation Server for phasing and imputation analyses [14], while those from dbGaP cohorts was imputed with 1000 genome project Phase 3 reference panel[15]. Default parameters were utilized for the imputation process.

Following imputation, SNPs in the imputed datasets with an imputation score (R^2^) < 0.3 or a Minor Allele Frequency (MAF) < 1% were excluded from further analysis. An identity-by-state (IBS) matrix was calculated for each study and sample, and samples with an IBS similarity > 0.9 were excluded (as potential duplicates/first-degree relatives). Finally, clean sample sets for the three cohorts were retained for subsequent GWAS analyses.

#### GWAS summary statistics collection

Due to the lack of large-scale Chinese summary statistics to serve as the base dataset for PRS estimation, we utilized summary statistics derived from the UK Biobank (UKBB) and Psychiatric Genomics Consortium (PGC), restricted to individuals of European ancestry. The sample sizes of the corresponding GWAS are presented in **Table 2**. Our Chinese cohort 1 was used as the PRS target dataset, which underwent genotype imputation using 1000 genome phase3 reference panel and the same quality control protocol described previously.

**Table 2.** Sample size of GWAS summary statistics after QC.

| Traits from UK Biobank | Sample size (EUR) of summary statistics |
| --- | --- |
| IQ | 135,088 |
| PM | 419,951 |
| RT | 417,660 |
| MEM | 137,999 |

To study the shared genetic basis between three SCZ clinical phenotypes (aggressive acts, self-harm, and AAO) from cohort 1 (PRS target dataset) and other psychiatric/cognitive traits (PRS base dataset), we leveraged GWAS summary statistics for four psychiatric disorders: bipolar disorder (BIP), autism spectrum disorder(ASD), attention deficit hyperactivity disorder (ADHD), and major depressive disorder (MDD), downloaded from the PGC. In addition, to examine genetic overlap between SCZ clinical phenotypes and four cognitive functions — fluid intelligence score (IQ), prospective memory (PM), reaction time (RT), and memory (MEM), we used GWAS summary statistics for these cognitive phenotypes downloaded from UKBB, as detailed in Table 1.

### Multiple testing control

We employed a false discovery rate (FDR) to control for multiple tests in post-GWAS analyses. FDR controls the expected proportion of false positives among variants declared significant. The traditional Bonferroni correction imposes an increasingly stringent threshold when more hypotheses are tested, which tends to be over-conservative in high-throughput studies.

### Genome-wide association study on clinical phenotype and meta-analysis of GWAS results

GWAS analyses were initially performed for each cohort individually. Logistic regression was employed for binary traits, while linear regression was used for continuous traits. Several variables, including sex, age, and the top four genetic principal components, were included as covariates. Notably, the age covariate was excluded from the GWAS analysis for the AAO trait to minimize the correlation with the AAO itself. The genomic control parameter (λ_GC_) was calculated for each analysis to assess the potential inflation of test statistics due to residual population stratification. A genomic control λ ranging from 0.9 to 1.1 indicates no obvious inflation of test statistics from uncorrected population stratification or other systematic biases. The conventional thresholds of *P* < 5E−08 and *P* < 5E−06 were considered as genome-wide significant and suggestive thresholds, respectively. Genomic risk loci were identified by clustering SNPs that exhibited high linkage disequilibrium (LD) with the top significant SNPs (LD r^2^ > 0.3, distance = 500kb).

After the individual GWAS analysis for each cohort, a meta-analysis was conducted for different combination of cohorts using the METAL program [16]. GWAS summary statistics, sharing the same trait from the cohorts were meta-analysed using an inverse variance–weighted fixed-effects model implemented in the program. Specifically, meta-analyses were conducted to analyse the history of self-harm in the two cohorts, as well as the history of aggressive acts in cohort 1 and violent acts in cohort 2. Additionally, the AAO phenotype was meta-analysed across all three cohorts. The genome-wide significant and suggestive thresholds remained the same as mentioned above.

### Deciphering the genetic relationship of SCZ phenotypes with psychiatric disorders/traits

#### PRS Calculation

PRSice-2[17], specialized software for PRS calculation in large-scale genotype-phenotype datasets, was utilized to estimate the PRS for the four psychiatric disorders and four cognitive traits. The software selected the optimal p-value threshold, maximizing the variance explained by the SNPs with p-values below the selected threshold. LD clumping was performed using an *R^2^* threshold of 0.15 and a window size of 250 kb to account for the correlation structure between SNPs.

### Genome-wide gene-based association analysis and transcriptomics-wide association study (TWAS) analysis

#### Identification of significant genes and enriched pathways for SCZ phenotypes

To elucidate the underlying genetic basis of the GWAS and meta-analysis summary statistics, we employed two different approaches, MAGMA and MetaXcan, to map associated SNPs to their corresponding associated genes. The MAGMA program utilized the original p-values from the SNP-based GWAS analysis to identify candidate genes associated with a trait. By aggregating the statistical significance of SNPs within a gene, the program generated gene-based statistics [18]. Genes with a p-value below 0.05 were deemed nominally associated with each trait.

In the second approach, MetaXcan integrates genomic information from cis-eQTLs (expression quantitative trait loci) derived from GTEx with GWAS summary-level data to identify the genes associated with a trait, eliminating the need for individual-level genotyping data [19]. MetaXcan consists of two main programs. The S-PrediXcan program computes associations for single tissues, while the latter program computes associations for multiple tissues and integrates measurements across tissues, including 13 subtypes of brain tissue. By conducting multiple S-PrediXcan analyses on various tissues, the program successfully mitigates the false negative rate induced by the stringent Bonferroni correction [20]. Genes with a p-value below 0.05 were considered nominally associated with each trait in the brain tissue.

#### Gene set and pathway enrichment analysis for trait-specific genes

To investigate the molecular mechanisms in which the identified genes may be involved in SCZ-related traits, we conducted Gene Ontology (GO) sets and Kyoto Encyclopedia of Genes and Genomes (KEGG) pathways enrichment analyses. These analyses were performed using the R package ‘ClusterProfiler’ [21]. The genes with a p-value below 0.05 were used for the enrichment analysis. For each set of genes, we examined the enrichment of GO sets and KEGG pathways. The enriched GO sets or KEGG pathways with a p-value below 0.05 were considered statistically significant, indicating their potential involvement in the molecular mechanisms underlying SCZ-related traits.

### Prediction of severity in SCZ patients

We included a subset of our HK cohort (N=669) that have both the genotype data and related phenotypes for building these prediction models.Among those recruited subjects, there were totally 275 cases (with ≥ 1 hospital admission record due to SCZ) and 394 controls (without any SCZ-related hospital admission records). The number of hospitalizations for SCZ patients is shown in **Table 3**, while **Figure 2** corresponds to the distribution of their total duration of hospitalization.

**Figure 2.**
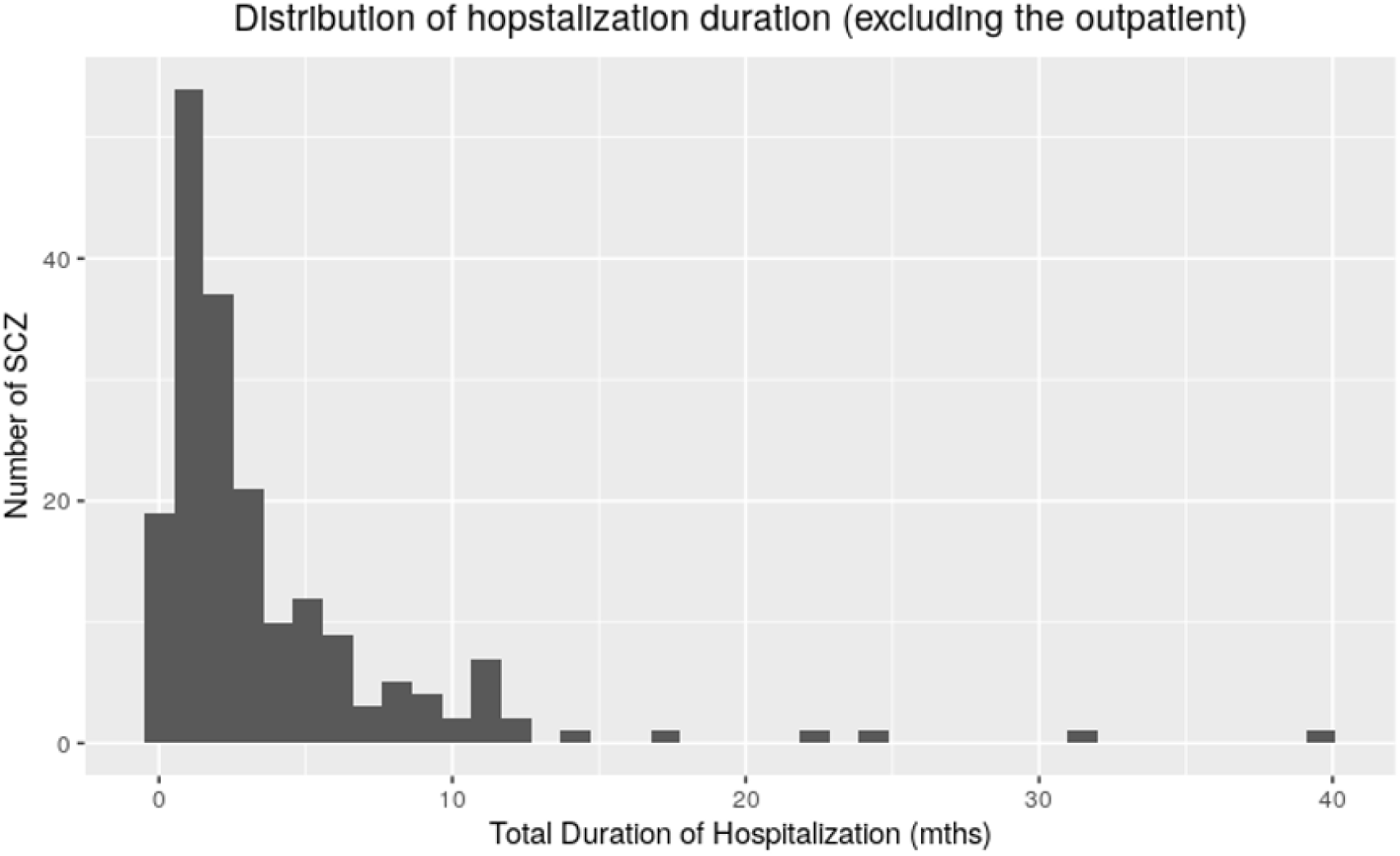
Histogram of the hospitalization duration for cohort 1 (without outpatient)

**Table 3.** Distribution of number of hospitalizations for SCZ patients.

| **No. of Hospital Admissions** | 0 | 1 | 2 | 3 | 4 | 5 | 6 | 7 | 8 |
| --- | --- | --- | --- | --- | --- | --- | --- | --- | --- |
| **No. of SCZ patients** | 394 | 162 | 59 | 30 | 16 | 5 | 1 | 1 | 1 |

The model fitting procedure follows a 5-fold cross-validation design by randomly dividing these 669 subjects into training and testing set in a ratio of 80:20. The PRS scores for the subjects in the training and testing set were calculated by PRSice-2[17]. To avoid overfitting issues, optimal P-value threshold auto-detected by PRSice-2 was only used for calculating the training set PRS score, where the PRS estimation of testing set was based on the same P-value threshold derived from the training set PRS calculation.

A highly-optimized gradient boosting ML model was built with the training dataset by implementing the R package XGBoost[22]. With the predictors extracted from the testing set, we could predict different outcomes with this XGBoost model. For the binary outcome: hospitalization or not, the accuracy of prediction model to discriminate case and control was assessed using the area under the receiver operator characteristic curve (ROC-AUC); While for the number and total duration of hospitalization, the prediction performance was measured by the variance explained (R^2^) by training model and root mean squared error (RMSE) between the predicted and true value of the corresponding outcomes. After repeating the above steps five times for cross-validation, we derived the final model performance metric ROC-AUC or R^2^ and RMSE by averaging the 5-fold cross-validation results.

To explain the prediction made by our ML models, we further conducted risk factor analyses by Shapley values, a concept from cooperative game theory introduced in the 1950s. With the implementation of R package SHAPforxgboost, we can quantify the marginal contribution of not only each single predictor, but also their interactions to the severity outcomes of SCZ[23].

## Results

### Genome-wide association study on clinical phenotype and meta-analysis of GWAS results

#### SCZ cohort 1

For the SCZ cohort 1 patients, after applying stringent quality controls, a total of 677 patients were retained for further analyses. In the GWAS analyses for five phenotypes (a history of aggressive acts, a history of self-harm, hospitalization-Yes/No, hospitalization count, AAO), we identified one genome-wide significant variant associated with hospitalization count on chromosome 9 (rs60648049, *P* = 1.05E-08, **Table 4 and Table ST5;** QQ plot and Manhattan plot in **Figure 3**). Furthermore, a clumping analysis was performed within a 500kb distance of the genomic location surrounding the observed variant. This analysis revealed nine SNPs that showed significant correlation with the variant (**Table ST1**). These findings suggest that the top variant is an independently associated SNP with the hospitalization count. The number of SNP signals that passed the suggestive threshold for the five phenotypes ranged from 9 to 111 (GWAS_hospitalization-count_ = 111, GWAS_AAO_ = 59, GWAS_self-harm_ = 50, GWAS_hospitalization_ = 9, GWAS_aggressive-acts_ = 9; **Table 4 and Table ST5**). The genomic inflation (λ_GC_) values for all five traits were within the range of 0.9 to 1.1, indicating no significant inflation of test statistics from uncorrected population stratification or other systematic biases.

**Figure 3.**
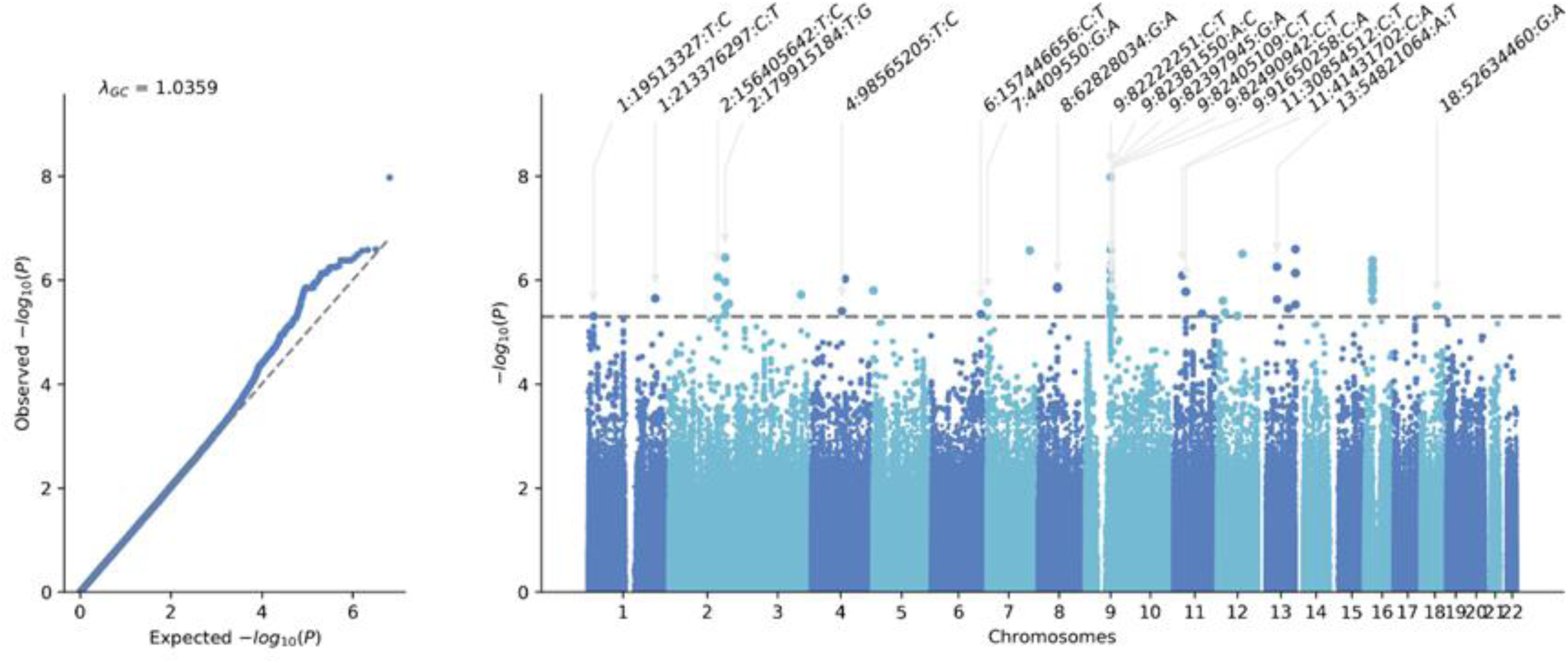
QQ plot and Manhattan plot of hospitalization count in SCZ cohort 1.

**Table 4.** GWAS analyses for target clinical phenotypes in three SCZ cohorts.

| **Cohort** | **Phenotypes** | **Ncase** | **Ncontrol** | **No. of SNPs**<br>**(*P* < 5E-08)** | **No. of SNPs**<br>**(*P* < 5E-06)** | **No. of SNPs**<br>**(*P* < 5E-03)** |
| --- | --- | --- | --- | --- | --- | --- |
| 1 | Aggressive acts | 138 | 537 | 0 | 9 | 27,202 |
| 1 | Hospitalization-Yes/No | 283 | 393 | 0 | 9 | 28,436 |
| 1 | A history of self_harm | 131 | 543 | 0 | 50 | 26,787 |
| 1 | Age at onset | N/A | N/A | 0 | 59 | 38,021 |
| 1 | Hospitalization count | N/A | N/A | **1** | 111 | 34,875 |
| 2 | Age at onset | N/A | N/A | **1** | 48 | 32,149 |
| 2 | A history of self_harm | 125 | 261 | 0 | 1 | 25,705 |
| 2 | Violent acts | 217 | 169 | 0 | 48 | 25,788 |
| 2 | Patterns of SCZ episodes | 83 | 268 | 0 | 11 | 25,088 |
| dbGaP | Age at onset | N/A | N/A | **2** | 208 | 74844 |

#### SCZ cohort 2

For the SCZ cohort 2 patients, a total of 387 subjects were included in the subsequent GWAS analyses. We identified one variant that passed the genome-wide significant threshold in the GWAS analysis with AAO (rs142498233, *P* = 1.20E-08, **Table 4**; QQ plot and Manhattan plot in **Figure 4**). In addition, the clumping analysis identified three SNPs that exhibited significant correlation with the variant. This analysis further supports the notion that the top variant is an independent associated SNP with the AAO phenotype (**Table ST2**). The number of SNP signals that passed the suggestive threshold for the four traits ranged from 1 to 48 (GWAS_AAO_ = 48, GWAS_violent-acts_ = 48, GWAS_episode_ = 11, and GWAS_self-harm_ = 1, **Table 4 and Table ST5**). The genomic inflation (λ_GC_) values for all four traits were within the range of 0.9 to 1.1, indicating no obvious inflation of test statistics.

**Figure 4.**
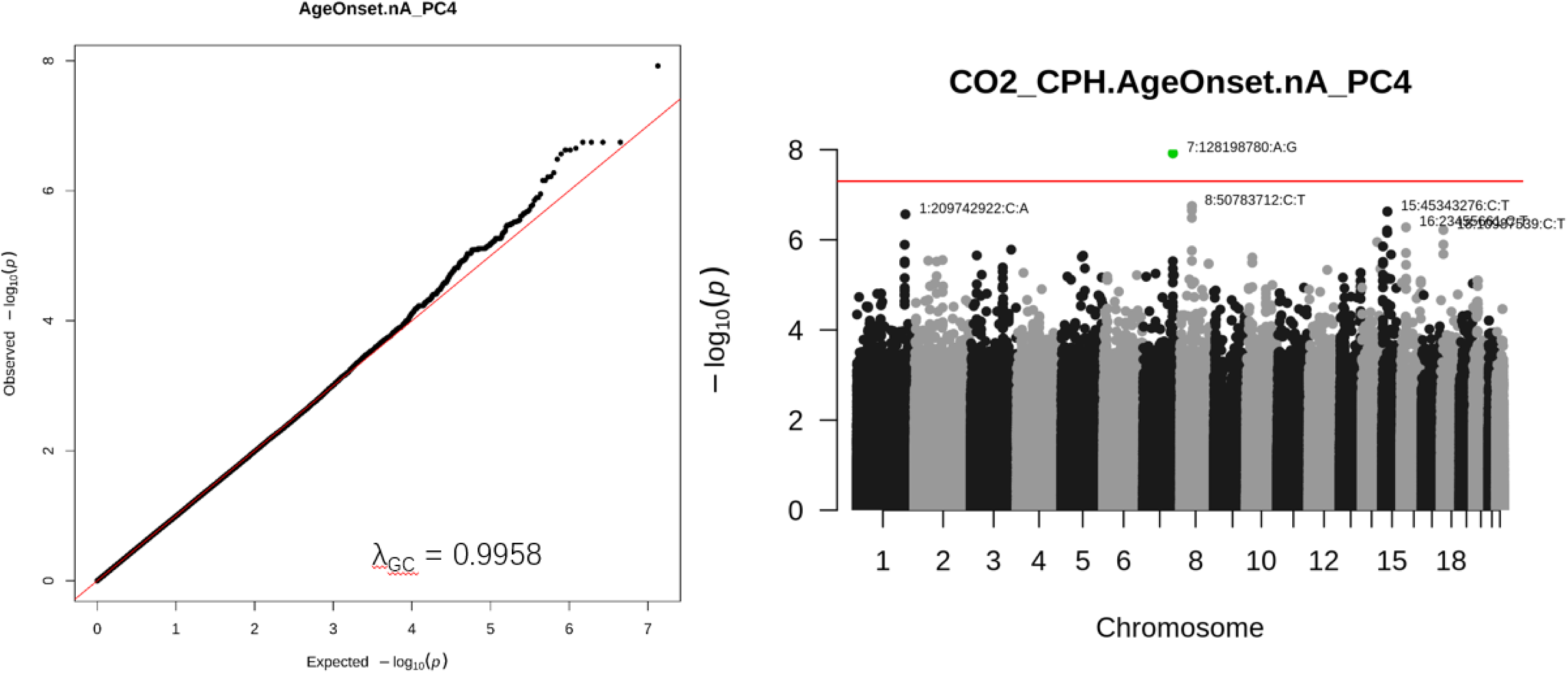
QQ plot and Manhattan plot of Age at onset in SCZ cohort 2.

#### dbGaP cohort

For the dbGaP cohort, 274 subjects and 845,780 SNPs survived after applying the quality control protocols. In the follow-up GWAS analyses, we found two SNP signals significantly associated with AAO in SCZ patients ( rs185213255, *P* = 4.17E-08; rs144645158, *P* = 4.56E-08; **Table 4 and Table ST5**; QQ plot and Manhattan plot in **Figure 5**). Furthermore, the clumping analysis revealed 8 SNPs significantly correlated with the variants rs185213255, and 37 SNPs significantly correlated with the variant rs144645158 (**Table ST3**). Moreover, a total of 208 SNPs surpassed the genome-wide suggestive threshold (**Table 4 and Table ST5**). To assess potential genomic inflation, the λ_GC_ value was calculated to be 1.007, indicating no significant inflation of test statistics.

**Figure 5.**
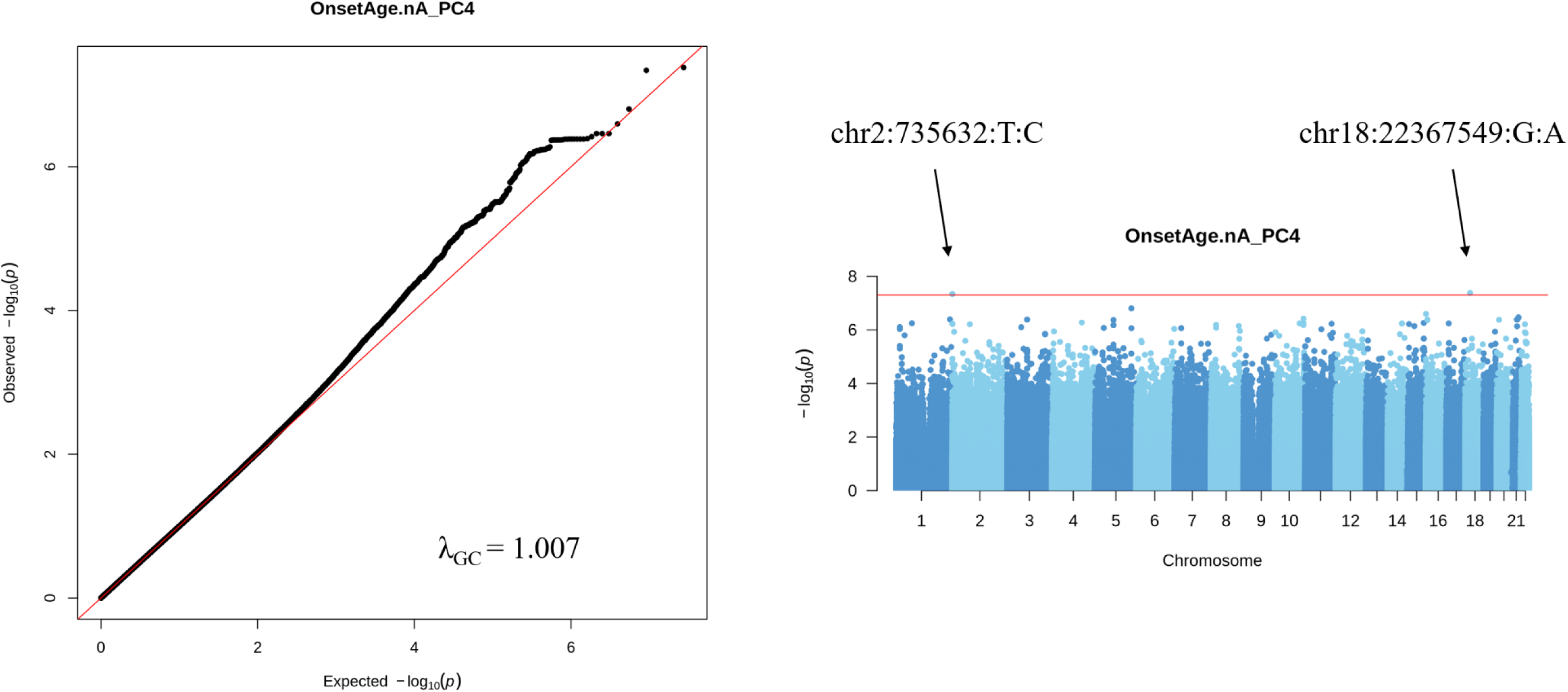
QQ plot and Manhattan plot of Age at onset in dbGaP SCZ cohort.

#### Meta-analyses for the overlapping phenotypes across the three cohorts

To increase the statistical power and identify true positive SNP signals, we conducted meta-analyses for the overlapping phenotypes across the three different cohorts (AAO). In the meta-analysis results, three SNPs were found to be significantly associated with AAO in SCZ patients ( rs142498233, *P* = 5.66E-09; rs185213255, *P* = 1.61E-08; rs144645158, *P* = 1.77E-08; **Table 5 and Table ST5;** QQ plot and Manhattan plot in **Figure 6**). The variant rs142498233 was also identified in cohort 2, and variants rs185213255 and rs144645158 were consistent with the variants identified in the dbGaP cohort. Furthermore, a total of 301 SNPs surpassed the genome-wide suggestive threshold (**Table ST5**). The correlated SNPs around these three SNPs are shown in **Table ST4**. It is worth noting that the meta-analysis results did not indicate any significant genomic inflation of test statistics, ensuring the reliability of the findings.

**Figure 6.**
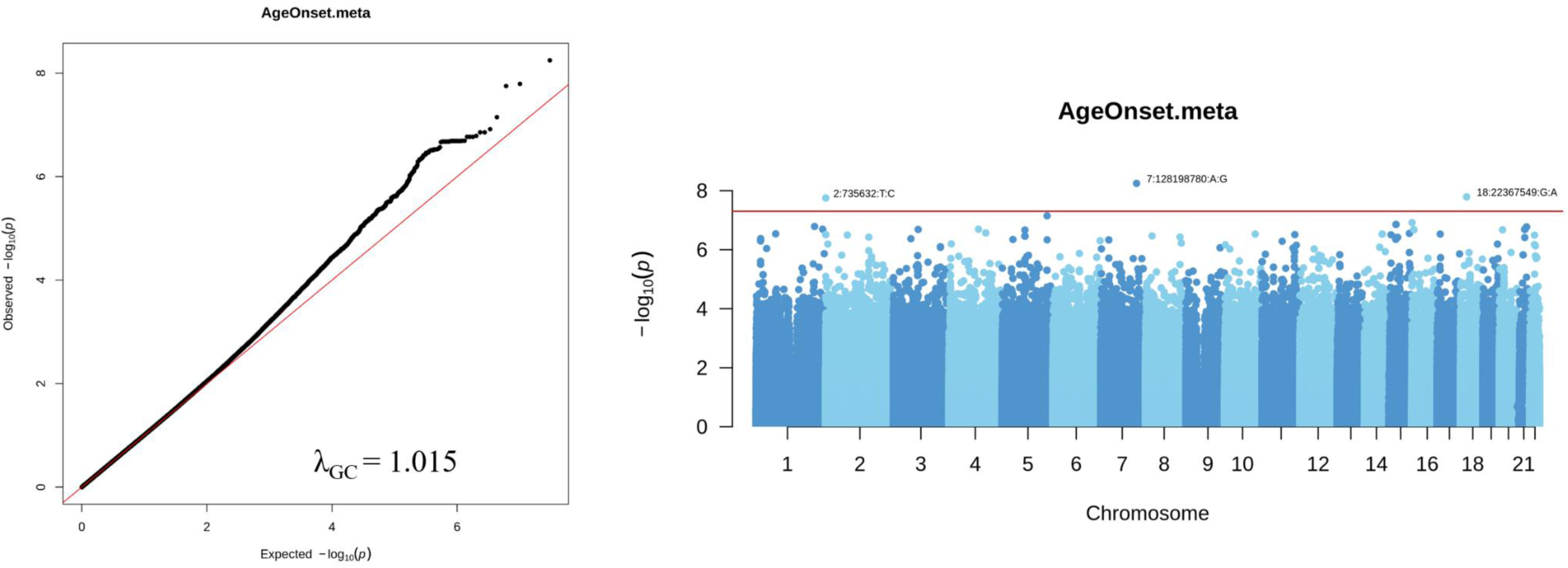
QQ plot and manhattan plot for the meta-analysis of Age at onset across the three cohorts.

**Table 5.** Meta-analyses for the overlapping phenotypes across the three cohorts.

| Cohorts | Phenotypes | nSNP | $P < 5E-08$ | $P < 5E-06$ | $P < 5E-03$ | $\lambda_{GC}$ |
| --- | --- | --- | --- | --- | --- | --- |
| Three cohorts | AAO | 8,920,118 | 3 | 301 | 87,981 | 1.01454 |
| 1 and 2 | Self-harm | 6,883,578 | 0 | 6 | 27,840 | 0.98285 |
| 1 and 2 | Aggressive + violent acts | 6,891,781 | 0 | 21 | 29,427 | 0.99070 |

### Deciphering the genetic relationship of SCZ phenotypes with psychiatric disorders/traits

#### Aggressive Behaviours

After adjusting for gender and education level, our analyses provided significant insights into the behavioural phenotypes under investigation. The adjusted models revealed a distinct severity in aggressive behaviours among males and individuals with SCZ who had lower education level (*P* = 0.003; *P* = 0.018). Additionally, there was a strong association between aggressive acts and a diminished PRS for IQ, contrasted with an elevated PRS for MEM (*P* = 0.032; *P* = 0.015).

#### Self-Harm Behaviours

The tendency to engage in self-harm behaviours among the SCZ cohort demonstrated an increase in individuals possessing lower educational attainment levels, though the strength of this association was moderate (*P* = 0.099). Furthermore, an observable genetic association between self-harm behaviours and ADHD was identified, indicating a potential underlying genetic predisposition (*P* = 0.088).

#### Onset Age of SCZ

An earlier AAO of SCZ was predominantly observed among SCZ individuals possessing higher educational attainment and among males, with (P = 0.099). Additionally, younger AAO was associated with increased PRS for BIP and MEM within the SCZ cohort, as indicated by p-values of 0.090 and 0.060, respectively.

These exploratory results (**Table 6**), although not surviving correction for multiple-testing, highlight the complex interplay between genetic predispositions and socio-demographic factors in shaping behavioural phenotypes in individuals with SCZ.

**Table 6.**
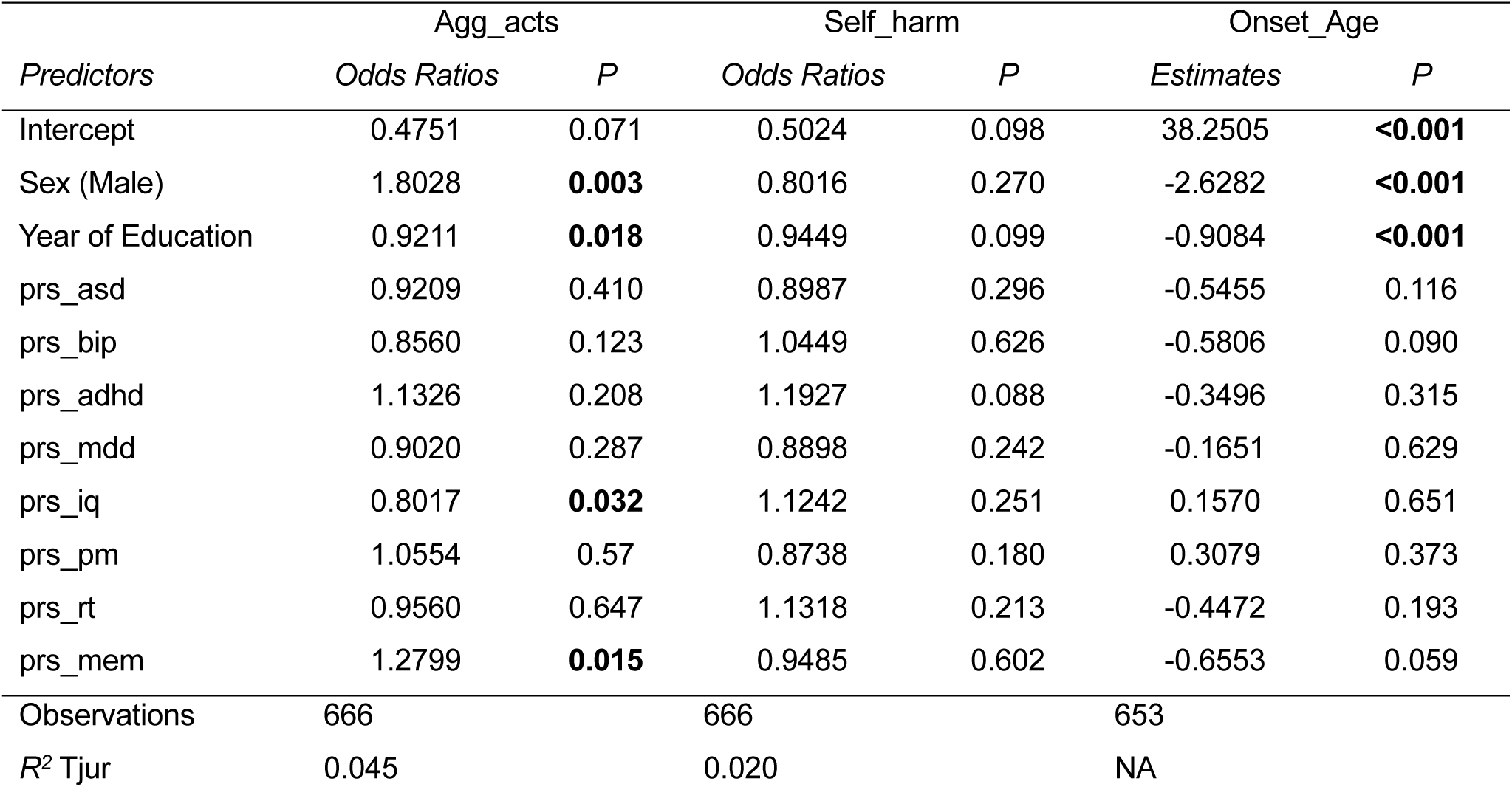
Shared genetic effect of psychiatry disorders and cognitive traits with the 3 SCZ phenotypes.

### Genome-wide gene-based association analysis and transcriptomics-wide association study (TWAS) analysis

#### Identification of significant genes and enriched pathways for SCZ phenotypes

To investigate the molecular pathogenesis of specific clinical phenotypes in SCZ patients, we employed the MAGMA and MetaXcan programs to conduct gene-based association analyses. The MAGMA program identified a range of 750 to 937 genes that exhibited nominal associations with multiple SCZ-related phenotypes, while no gene *achieved* a significant association with these phenotypes (**Table 7**). Subsequently, all nominally associated genes were subjected to Gene Ontology (GO) and Kyoto Encyclopedia of Genes and Genomes (KEGG) enrichment analyses for the respective clinical phenotypes (**Table 8** and **Table 9**).

**Table 7.** Summary of genome-wide gene-based association studies for clinical phenotypes in SCZ patients.

| Cohort | Phenotype | nGene | No. of Genes<br>($P < 0.05$) | No. of Genes<br>(FDR < 0.05) | No. of Genes<br>(FDR < 0.1) |
| --- | --- | --- | --- | --- | --- |
| 1 | AAO | 18,042 | 911 | 0 | 0 |
| 1 | Aggressive acts | 18,042 | 825 | 0 | 0 |
| 1 | Hospitalization count | 18,042 | 937 | 0 | 0 |
| 1 | Hospitalization | 18,042 | 930 | 0 | 0 |
| 1 | Self-harm | 18,038 | 802 | 0 | 0 |
| 2 | AAO | 18,037 | 868 | 0 | 0 |
| 2 | SCZ Episode | 18,027 | 809 | 0 | 0 |
| 2 | Self-harm | 18,035 | 750 | 0 | 0 |
| 2 | Violent acts | 18,037 | 891 | 0 | 0 |
| dbGaP | AAO | 18,939 | 935 | **0** | **0** |
| 1 and 2 | Self-harm | 17,857 | 802 | 0 | 0 |
| 1 and 2 | Aggressive + violent acts | 17,857 | 893 | 0 | 0 |
| Three cohorts | AAO | 17,857 | 866 | 0 | 0 |

Remark: nGenes: the total number of genes that analysed by the MAGMA program.

**Table 8.**
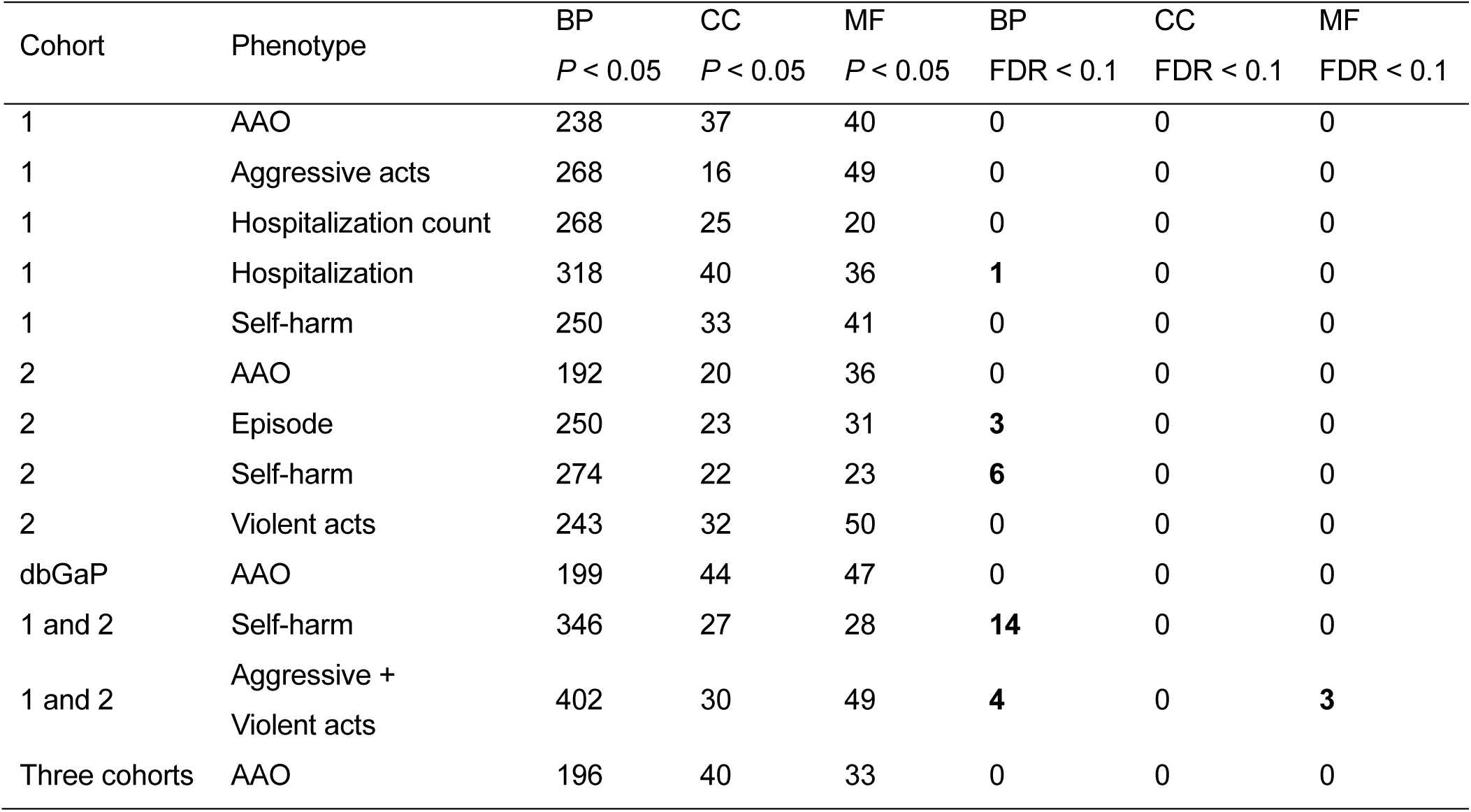
Summary of GO enrichment results for associated genes identified by MAGMA program.

| Cohort | Phenotype | BP | CC | MF | BP | CC | MF |
| --- | --- | --- | --- | --- | --- | --- | --- |
| | | $P < 0.05$ | $P < 0.05$ | $P < 0.05$ | FDR < 0.1 | FDR < 0.1 | FDR < 0.1 |
| 1 | AAO | 238 | 37 | 40 | 0 | 0 | 0 |
| 1 | Aggressive acts | 268 | 16 | 49 | 0 | 0 | 0 |
| 1 | Hospitalization count | 268 | 25 | 20 | 0 | 0 | 0 |
| 1 | Hospitalization | 318 | 40 | 36 | **1** | 0 | 0 |
| 1 | Self-harm | 250 | 33 | 41 | 0 | 0 | 0 |
| 2 | AAO | 192 | 20 | 36 | 0 | 0 | 0 |
| 2 | Episode | 250 | 23 | 31 | **3** | 0 | 0 |
| 2 | Self-harm | 274 | 22 | 23 | **6** | 0 | 0 |
| 2 | Violent acts | 243 | 32 | 50 | 0 | 0 | 0 |
| dbGaP | AAO | 199 | 44 | 47 | 0 | 0 | 0 |
| 1 and 2 | Self-harm | 346 | 27 | 28 | **14** | 0 | 0 |
| 1 and 2 | Aggressive +<br>Violent acts | 402 | 30 | 49 | **4** | 0 | **3** |
| Three cohorts | AAO | 196 | 40 | 33 | 0 | 0 | 0 |

**Table 9.** Summary of KEGG enrichment results for associated genes identified by MAGMA program.

| **Cohort** | **Phenotype** | **No. of Pathways**<br>***P* < 0.05** | **No. of Pathways**<br>**FDR < 0.1** |
| --- | --- | --- | --- |
| 1 | AAO | 10 | 0 |
| 1 | Aggressive acts | 7 | 0 |
| 1 | Hospitalization count | 5 | 0 |
| 1 | Hospitalization | 31 | **1** |
| 1 | Self-harm | 7 | 0 |
| 2 | AAO | 5 | 0 |
| 2 | SCZ Episode | 4 | 0 |
| 2 | Self-harm | 15 | 0 |
| 2 | Violent acts | 6 | 0 |
| dbGaP | AAO | 52 | 5 |
| 1 and 2 | Self-harm | 9 | 0 |
| 1 and 2 | Aggressive + Violent<br>acts | 17 | **3** |
| Three cohorts | AAO | 8 | 0 |

#### Gene set and pathway enrichment analysis for trait-specific genes

In the enrichment analyses, we observed that the nominally associated genes related to hospitalization were significantly enriched in the biological process of B cell activation involved in immune response (FDR = 2.10E-02, **Table 8** and **Table ST6**), as well as in the pathway associated with amoebiasis (FDR = 7.03E-02, **Table 9** and **Table ST7**). Moreover, the nominally associated genes linked to aggressive acts exhibited significant enrichment in various metabolic processes (GO gene sets) involving uronic acid, glucuronate, and flavonoid (FDR < 1.20E-02, **Table 8** and **Table ST6**). These genes were also significantly enriched in pathways associated with porphyrin metabolism (KEGG pathway database), including ascorbate and aldarate metabolism, and pentose and glucuronate interconversions (FDR < 2.01E-02, **Table 9** and **Table ST7**). These findings provide valuable insights into potential molecular mechanisms and biological processes contributing to the pathogenesis of the target phenotypes observed in SCZ patients.

For GO and KEGG enrichment analyses of the nominally associated genes, we observed that the genes associated with a history of self-harm in the GWAS analysis of cohort 1 were significantly enriched in the regulation of autophagy (FDR = 1.15E-02, **Table 11** and **Table ST9**). These genes also showed nominal associations with cellular components related to the outer membrane (FDR = 7.50E-02, **Table 11** and **Table ST9**). Furthermore, the genes associated with the AAO phenotype from the GWAS analysis of the dbGaP cohort exhibited significant enrichment in the molecular function of nuclease activity (FDR = 3.12E-02, **Table ST9**). Additionally, these genes demonstrated nominal associations with molecular functions involving catalytic activity acting on DNA (FDR = 7.53E-02) and ATP-dependent activity acting on DNA (FDR = 8.62E-02). However, no significant enriched signals were found in the KEGG enrichment analysis (**Table 12** and **Table ST10**).

**Table 10.** Summary of tissue-specific imputed gene expression association studies for clinical phenotypes in SCZ patients.

| Cohort | Phenotype | nGenes | No. of Genes<br>($P < 0.05$) | No. of Genes<br>(FDR < 0.1) |
| --- | --- | --- | --- | --- |
| 1 | Aggressive acts | 14089 | 514 | **7** |
| 1 | Hospitalization count | 14089 | 543 | 0 |
| 1 | Hospitalization | 14089 | 584 | **2** |
| 1 | AAO | 14089 | 573 | 0 |
| 1 | Self-harm | 14089 | 520 | **8** |
| 2 | AAO | 14107 | 575 | **2** |
| 2 | Episode | 14107 | 565 | 0 |
| 2 | Self-harm | 14107 | 531 | **2** |
| 2 | Violent acts | 14107 | 600 | **1** |
| dbGaP | AAO | 14205 | 439 | **3** |
| 1 and 2 | Self-harm | 14113 | 486 | **1** |
| 1 and 2 | Aggressive + Violent<br>acts | 14113 | 512 | **1** |
| Three cohorts | AAO | 14205 | 497 | 0 |

nGenes: number of analysed genes by the S-MultiXcan program in the whole brain tissue.

**Table 11.**
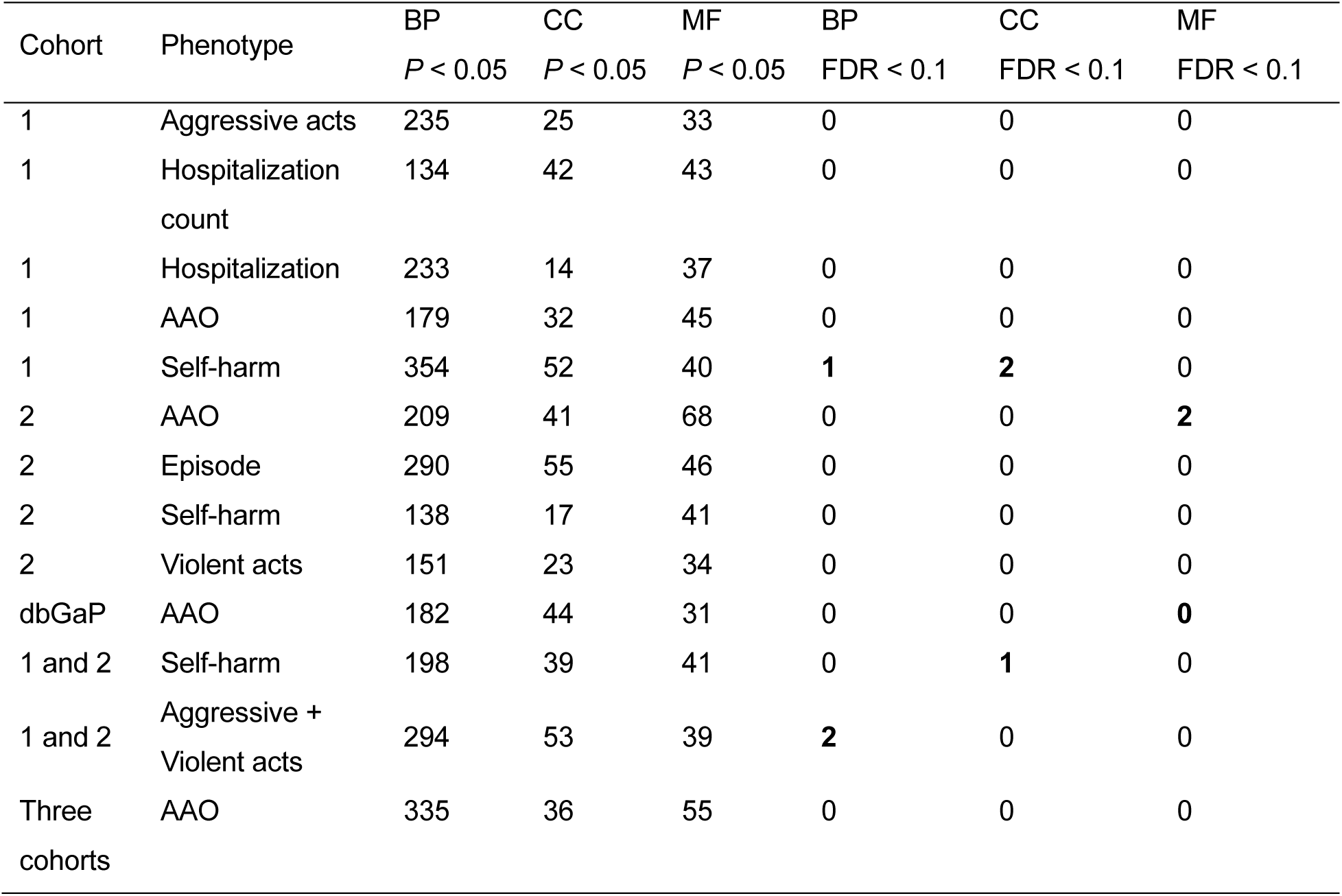
Summary of GO enrichment results for associated genes identified by S-MultiXcan program.

**Table 12.** Summary of KEGG enrichment results for associated genes identified by S-MultiXcan program.

| Cohort | Phenotype | No. of Pathways | No. of Pathways |
| --- | --- | --- | --- |
| | | $P < 0.05$ | FDR < 0.1 |
| 1 | Aggressive acts | 22 | 0 |
| 1 | Hospitalization count | 11 | 0 |
| 1 | Hospitalization | 8 | 0 |
| 1 | AAO | 28 | 0 |
| 1 | Self-harm | 11 | 0 |
| 2 | AAO | 7 | 0 |
| 2 | Episode | 26 | 0 |
| 2 | Self-harm | 6 | 0 |
| 2 | Violent acts | 9 | 0 |
| dbGaP | AAO | 13 | 0 |
| 1 and 2 | Self-harm | 9 | 0 |
| 1 and 2 | Aggressive + violent acts | 34 | 0 |
| Three cohorts | AAO | 37 | 2 |

#### Identification of significant genes in brain tissue by TWAS and enriched pathways for SCZ phenotypes

To investigate the potential association between genetically imputed gene expression and SCZ-related phenotypes at a tissue-specific level, we utilized the S-PrediXcan and S-MultiXcan programs to perform the gene-based association analyses. In the target brain tissue, this program identified a range of 469 to 600 genes that exhibited nominal associations with multiple SCZ-related phenotypes (**Table 10** and **Table ST8**). Notably, for aggressive acts in cohort 1, the *RIC8A* (resistance to inhibitors of cholinesterase 8 homolog A) gene displayed significant association in the putamen-basal-ganglia region (FDR = 2.33E-05, **Table ST8**). This association was further confirmed through meta-analysis of aggressive acts in the same region (FDR = 1.73E-04, **Table ST8**). Additionally, the *DDX58* (DEAD Box Protein 58, also called RIGI) gene showed associations with a history of self-harm in the cerebellar hemisphere for both cohort 1 (FDR = 6.91E-02) and cohort 2 (FDR = 1.61E-02, **Table ST8**). Regarding the AAO phenotype, several significant signals were identified, including the *DDIAS* (DNA damage induced apoptosis suppressor) gene in the frontal cortex region (FDR = 1.38E-3), the *GAS2* gene (FDR = 8.08E-3), and the *NBPF26* (neuroblastoma breakpoint family member 26) gene (FDR = 1.37E-03) in the cerebellum region (**Table ST8**). Moreover, the *ADGRE2* (adhesion G protein-coupled receptor E2) gene demonstrated significant association with hospitalization in the amygdala region (FDR = 2.55E-02, **Table ST8**).

### Prediction of severity in schizophrenia patients

#### Prediction of hospitalization or not

Represented by the ROC-AUC value, the 5-fold cross-validated prediction accuracy for the testing set is summarized in **Table 13**. The average ROC-AUC is 63.4% [59.1% - 67.8%], suggesting that a modest discrimination between cases and controls was achieved by our model. The risk ratio between the top 10% and bottom 10% percentile is approximately 2.5×, also indicating a modest discrimination power of hospitalization risk prediction. The variable importance (VarImp) plot for the XGBoost model is plotted in **Figure 7**, where AAO and aggressive acts were identified as the top 2 predictors with highest variable importance. While through the mean Shapley value overall VarImp plot as shown in **Figure 8**, we observed that lower AAO, having aggressive acts, higher PRS score of IQ and BIP, having self-harm acts were all associated with a higher mean Shapley value, suggesting their contributions to predict a higher risk of hospitalization. For interaction analysis, only the Shapley value interaction plots for the top 9 interaction pairs are shown in **Figure 9**. It was found that the combination of lower AAO and having aggressive acts contributes to a higher predicted hospitalization risk of SCZ, while however, higher AAO together with having aggressive acts in contrast lower this risk as indicated by the negative Shapley value. **Table 14** summarizes the permutation (N=500) P-value of the mean absolute Shapley value for different predictors, and only lower AAO, having aggressive and self-harm acts are statistically significant (P < 0.05) in association with higher risk of hospitalization.

**Figure 7.**
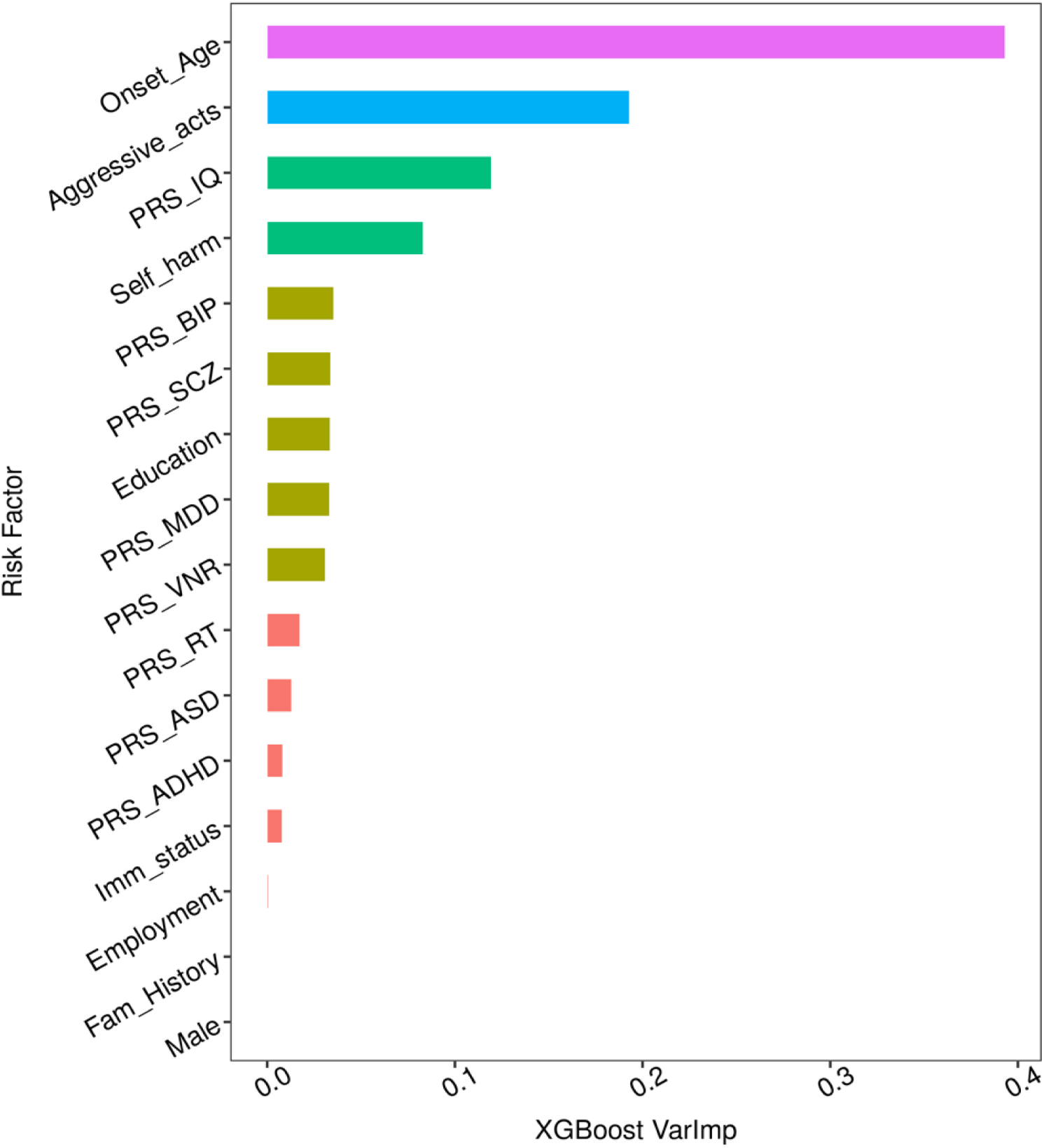
Variable importance plot of the XGBoost model for prediction of hospitalization or not.

**Figure 8.**
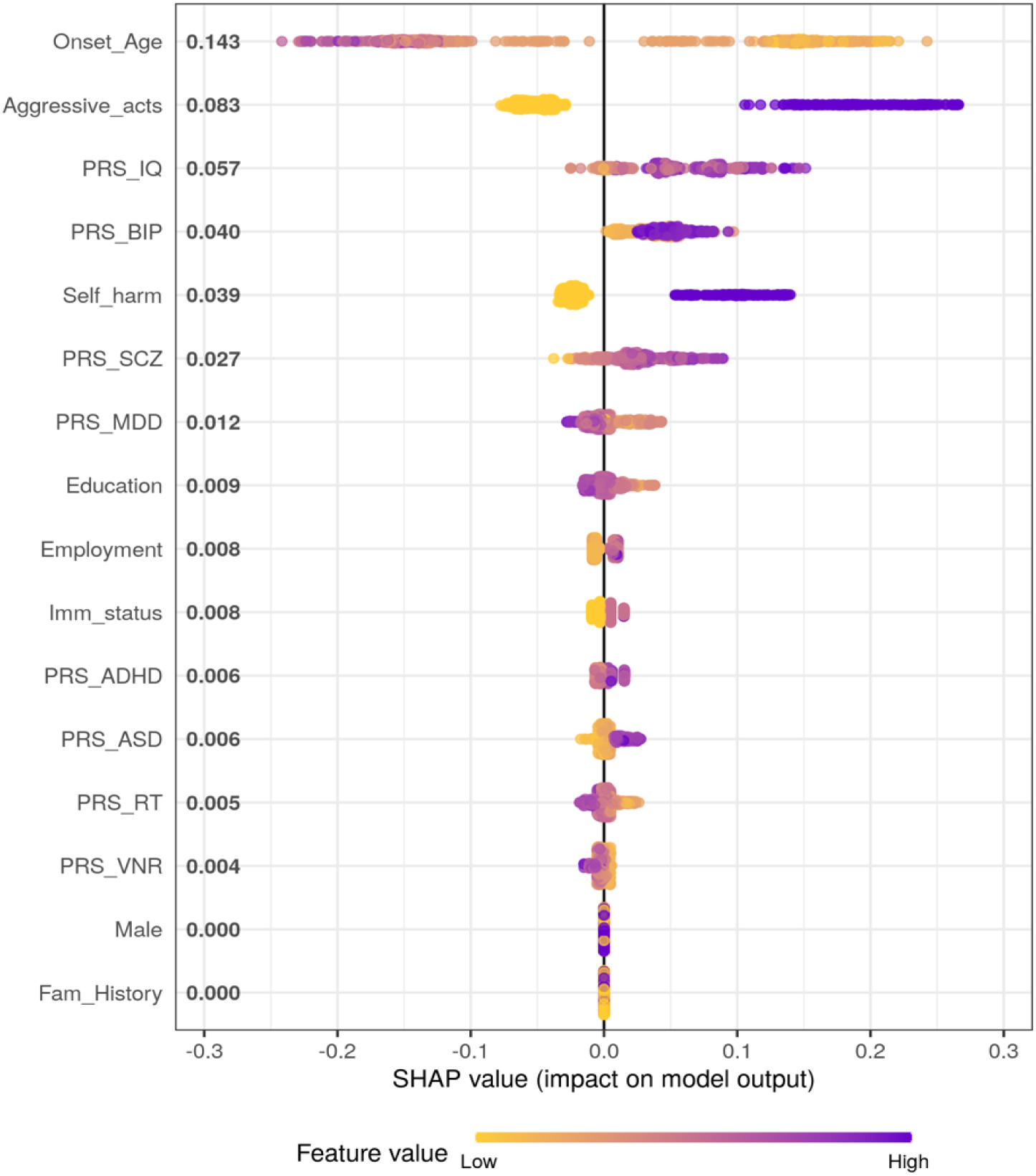
Mean Shapley value overall variable importance plot for prediction of hospitalization or not.

**Figure 9.**
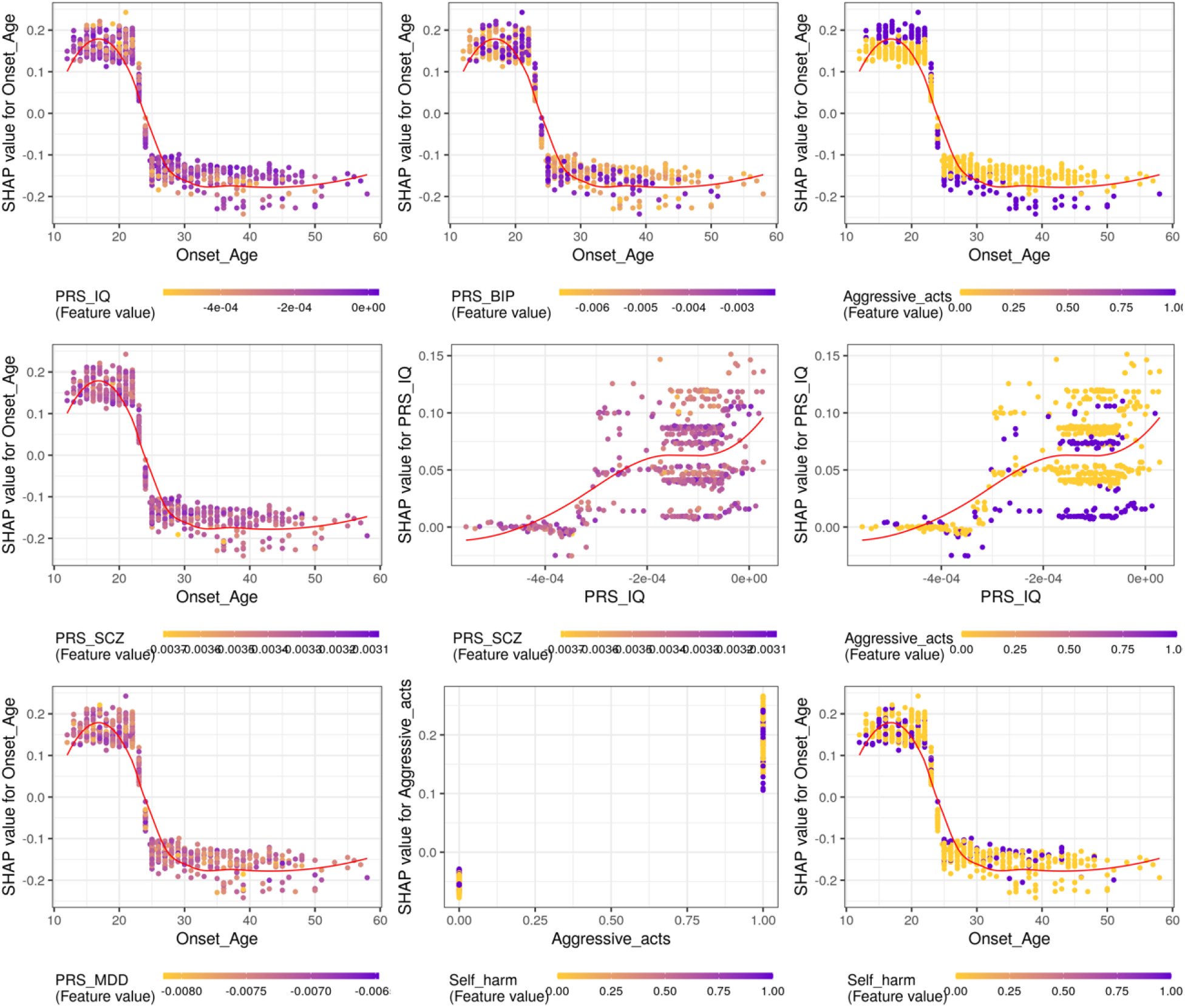
Shapley value interaction plot for the top 9 interaction pairs for prediction of hospitalization or not.

**Table 13.** Five-fold cross-validated prediction accuracy for prediction of hospitalization or not.

| **Fold** | **ROC-AUC** | **Standard Error for**<br>**ROC-AUC** | **Lower CI for ROC-**<br>**AUC** | **Upper CI for ROC-**<br>**AUC** |
| --- | --- | --- | --- | --- |
| **1** | 0.668 | 0.048 | 0.573 | 0.762 |
| **2** | 0.596 | 0.051 | 0.497 | 0.695 |
| **3** | 0.612 | 0.049 | 0.516 | 0.708 |
| **4** | 0.608 | 0.052 | 0.505 | 0.710 |
| **5** | 0.689 | 0.047 | 0.596 | 0.782 |
| **Average** | 0.634 | 0.022 | 0.591 | 0.678 |

**Table 14.**
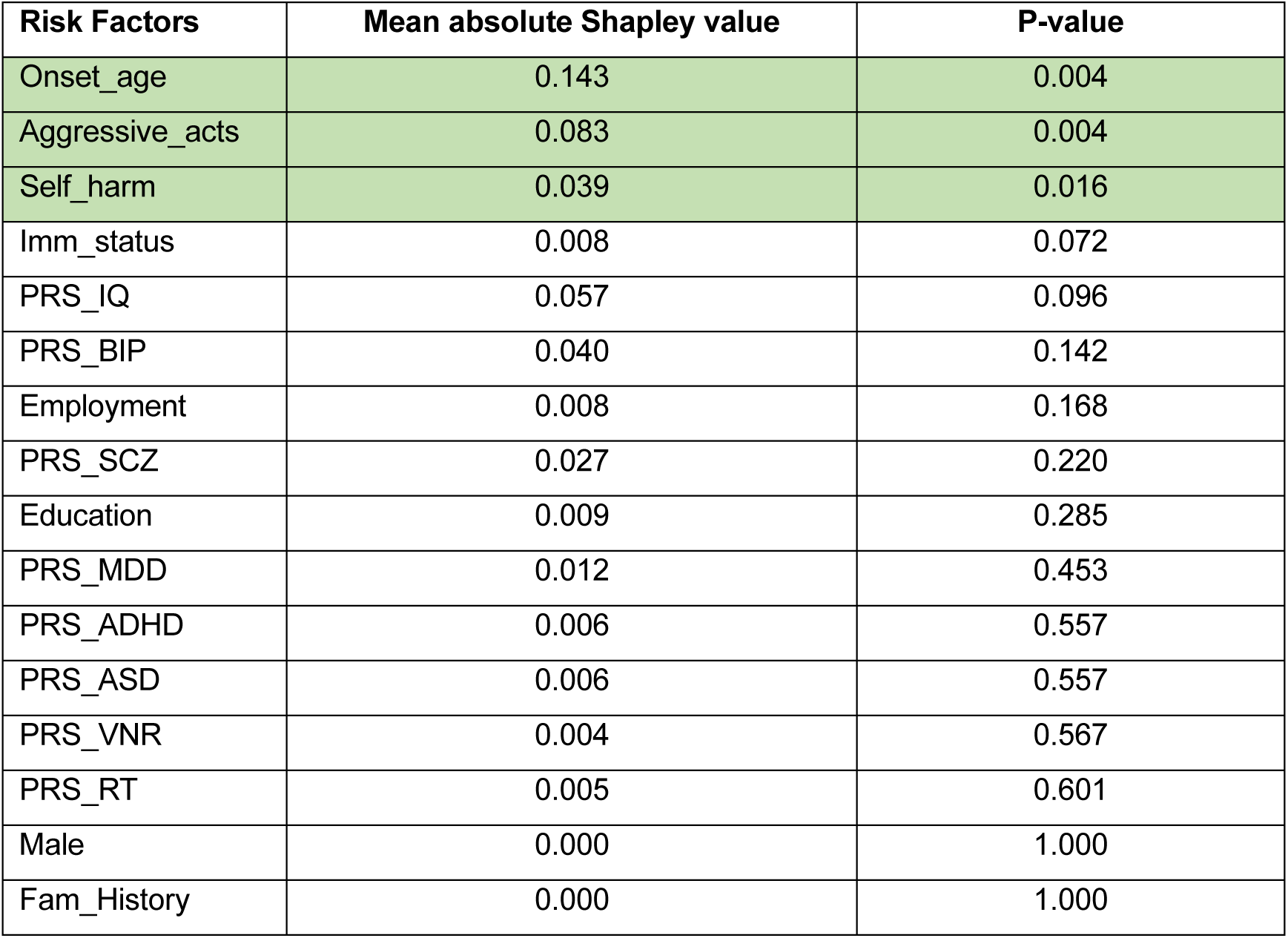
Permutation (N=500) P-value of mean absolute Shapley value for prediction of hospitalization or not.

#### Prediction of number of hospitalizations

The prediction performance as measured by RMSE and R^2^ is summarized in **Table 15**. The average RMSE of this 5-fold cross-validated model for predicting the number of hospitalizations is around 1.067, which is deemed acceptable. Additionally, this prediction model is also considered satisfactory by explaining about 8.9% of the variance on average. XGBoost VarImp plot is shown in **Figure 10**, with AAO and aggressive acts recognized as the top 2 predictors. Besides, by analyzing the mean Shapley values from the overall VarImp plot in **Figure 11**, we identified several factors contributing to the prediction of elevated number of hospitalizations, which includes: lower AAO, having aggressive and self-harm acts, higher PRS of BIP and lower education level. The interaction plot in **Figure 12** reveals a similar trend for lower AAO plus having aggressive acts to increase the number of hospitalizations, while the combination of higher AAO and having aggressive acts reverses this trend. Other combinations such as higher PRS for IQ with the presence of aggressive acts, and higher PRS for RT with engagement in self-harm acts also tend to elevate the SCZ severity via elevating the number of hospitalizations. As indicated in **Table 16**, only lower AAO, having aggressive and self-harm acts are statistically significant predictors (permutation P-value < 0.05) associating with the increased number of hospitalizations.

**Figure 10.**
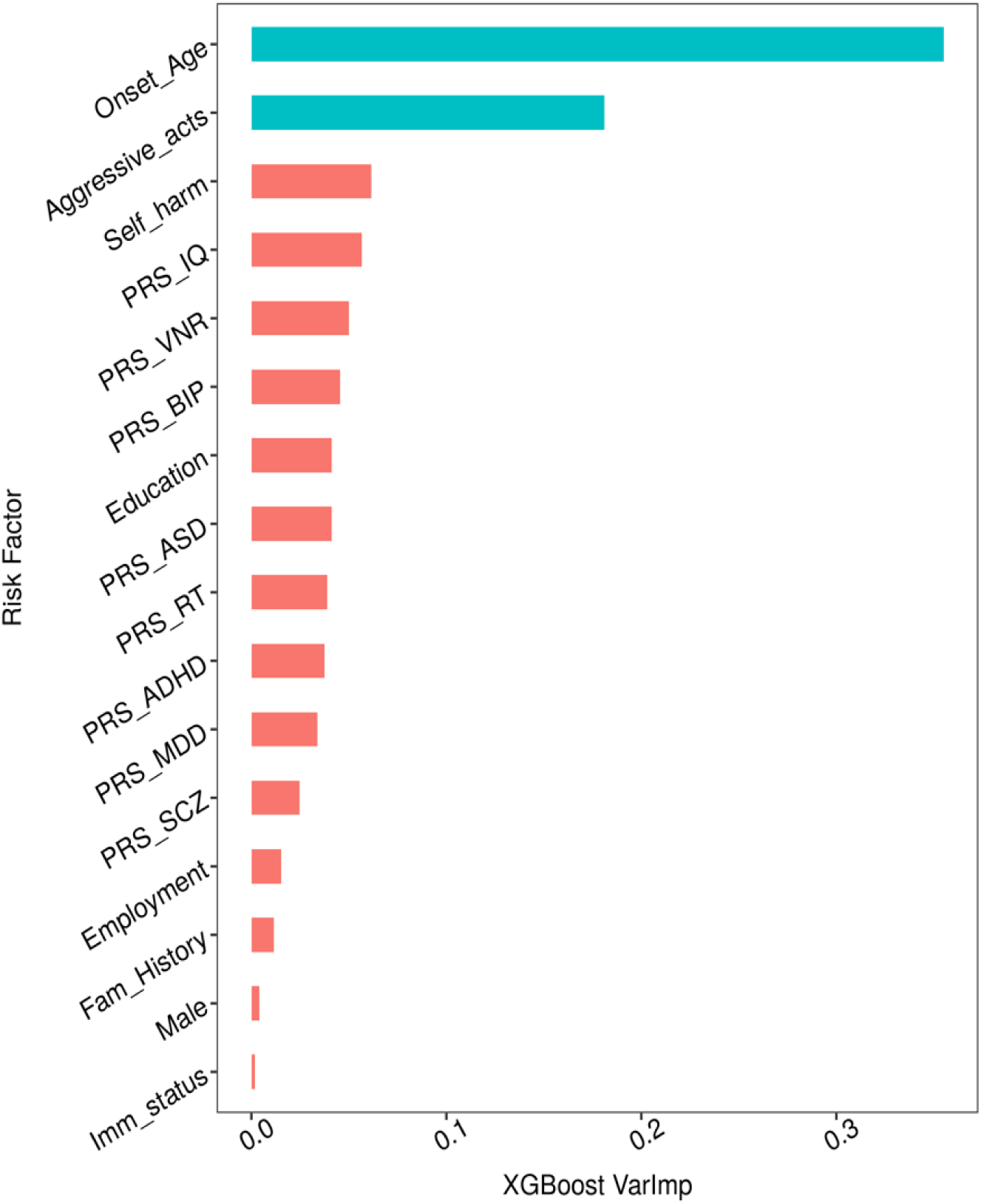
Variable importance plot of the XGBoost model for prediction of number of hospitalizations.

**Figure 11.**
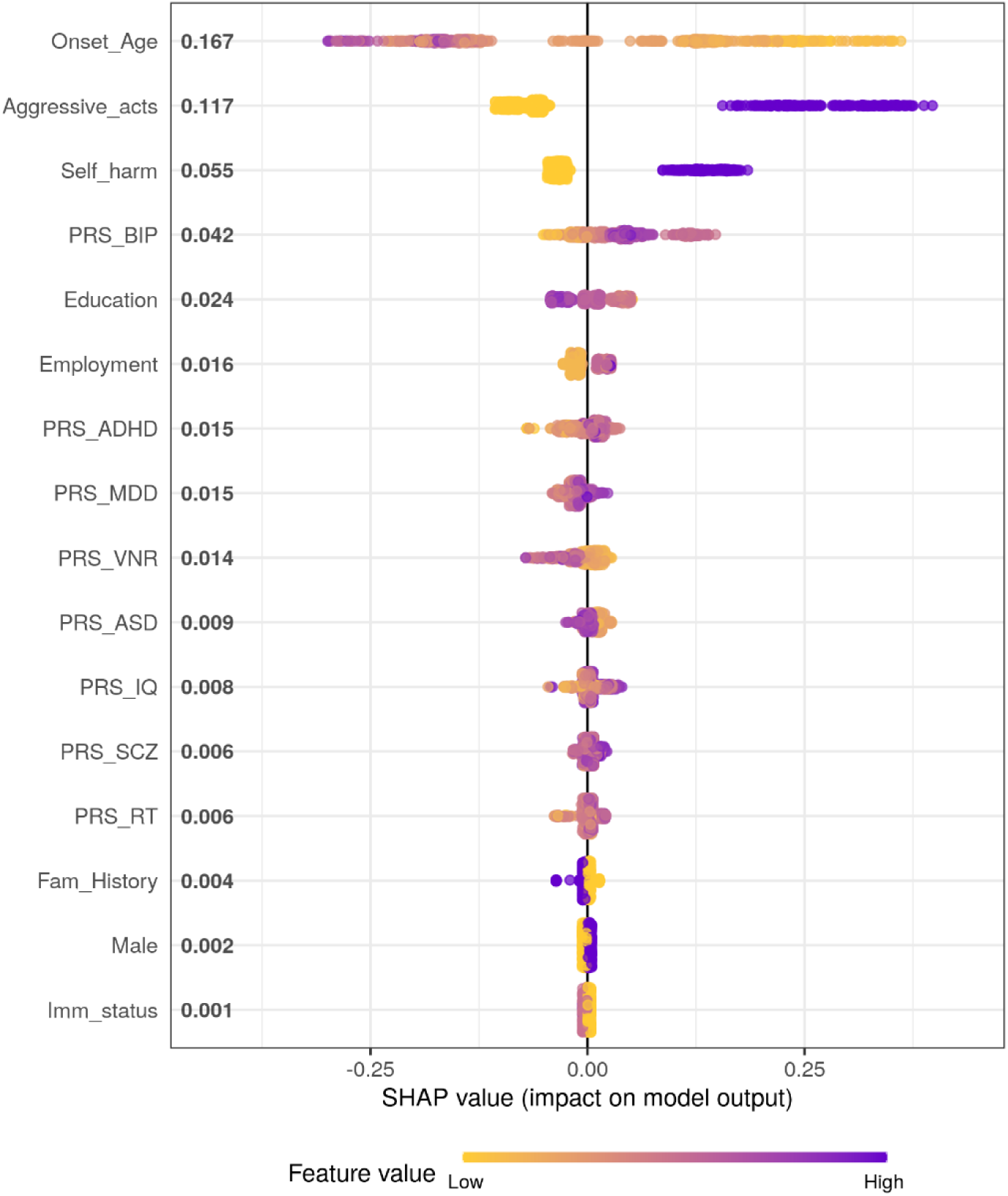
Mean Shapley value overall VarImp plot for prediction of number of hospitalizations.

**Figure 12.**
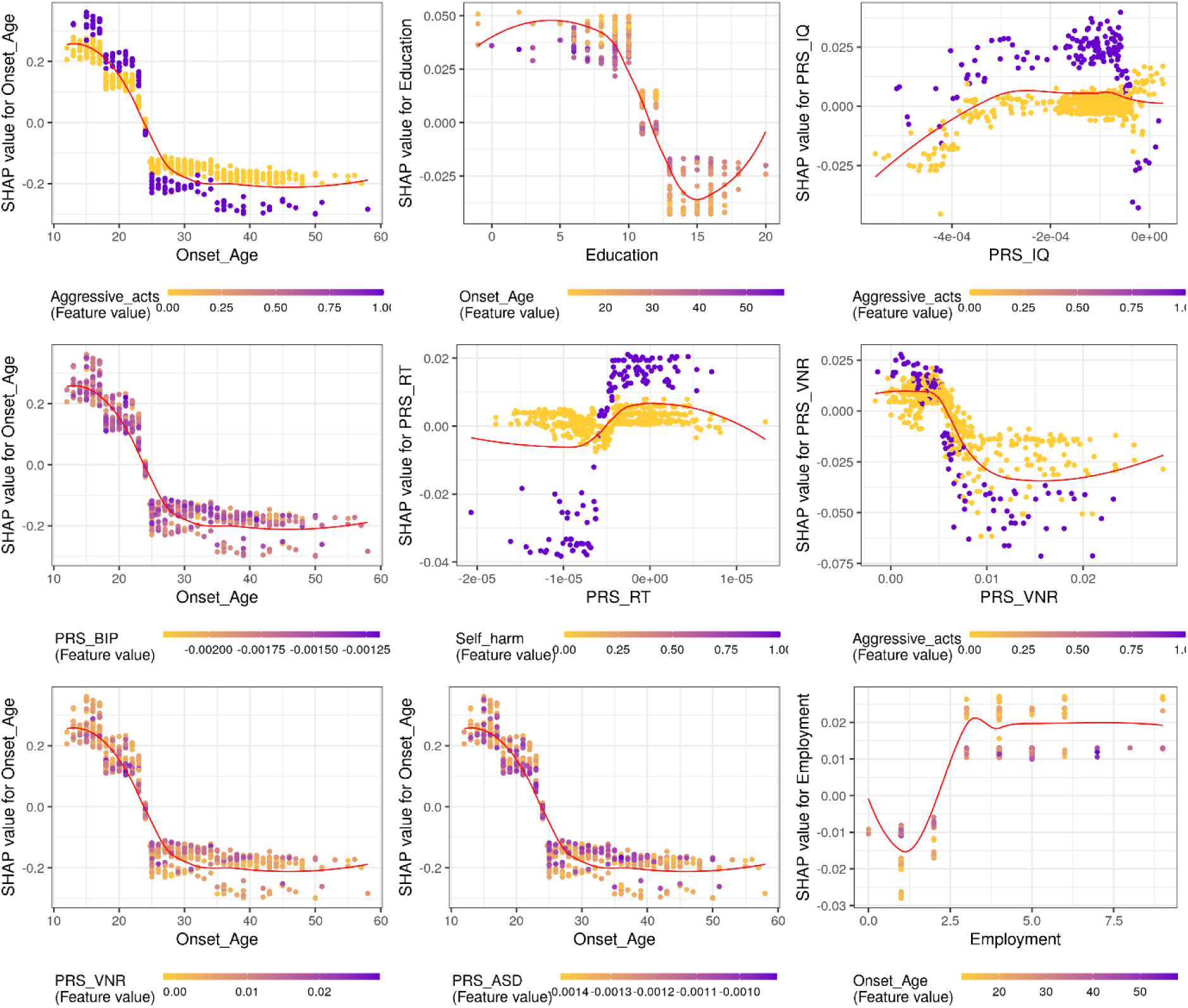
Shapley value interaction plot for the top 9 interaction pairs for prediction of number of hospitalizations.

**Table 15.** Five-fold cross-validated prediction accuracy for prediction of number of hospitalizations.

| **Fold** | **RMSE** | **R-Squared ($R^2$)** |
| --- | --- | --- |
| **1** | 1.145 | 0.095 |
| **2** | 0.934 | 0.094 |
| **3** | 1.252 | 0.054 |
| **4** | 1.060 | 0.097 |
| **5** | 0.942 | 0.106 |
| **Average** | 1.067 | 0.089 |

**Table 16.** Permutation (N=500) P-value of mean absolute Shapley value for prediction of number of hospitalizations.

| **Risk Factors** | **Mean absolute Shapley value** | **P-value** |
| --- | --- | --- |
| Onset_age | 0.167 | 0.001 |
| Aggressive_acts | 0.117 | 0.001 |
| Self_harm | 0.055 | 0.001 |
| PRS_BIP | 0.042 | 0.095 |
| Education | 0.024 | 0.112 |
| Employment | 0.016 | 0.125 |
| PRS_ADHD | 0.015 | 0.486 |
| PRS_MDD | 0.015 | 0.365 |
| PRS_VNR | 0.014 | 0.472 |
| PRS_ASD | 0.009 | 0.525 |
| PRS_IQ | 0.008 | 0.600 |
| PRS_SCZ | 0.006 | 0.650 |
| PRS_RT | 0.006 | 0.680 |
| Fam_History | 0.004 | 0.127 |
| Male | 0.002 | 0.164 |
| Imm_status | 0.001 | 0.196 |

#### Prediction of total duration of hospitalization

**Table 17** summarizes the model prediction accuracy. The 5-fold cross-validated RMSE is approximately 3.063 months, and the average R^2^ is around 2.8%, both suggesting an acceptable performance for the model to predict total duration of hospitalization. Aggressive acts and AAO are shown as the top 2 predictors from XGBoost VarImp plot in **Figure 13**. The mean Shapley value overall VarImp plot in **Figure 14** further identified several factors linked to the prediction of an increased duration of hospitalization, which involves: having aggressive acts, lower AAO, having self-harm acts and higher PRS for BIP. From the interaction analysis plot as shown in **Figure 15**, we observed that the combinations of lower AAO with either the presence of aggressive acts or a higher PRS for BIP were associated with an extended duration of hospitalization. Finally, regarding the permutation P-value, **Table 18** reveals that only having aggressive acts, lower AAO and having self-harm acts are the three factors significantly associated with the increase in total duration of hospitalization.

**Figure 13.**
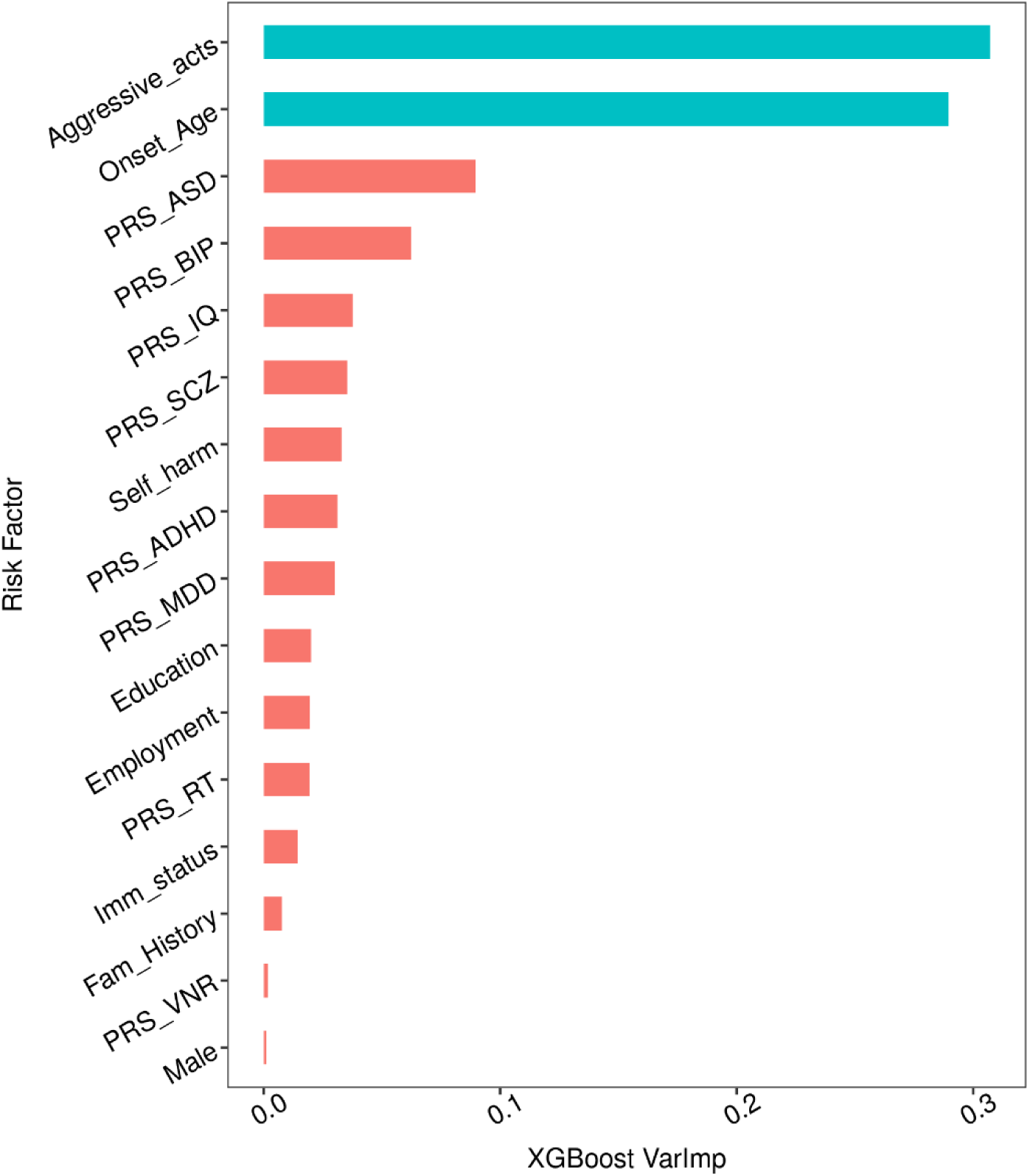
Variable important plot of the XGBoost model for prediction of total duration of hospitalization.

**Figure 14.**
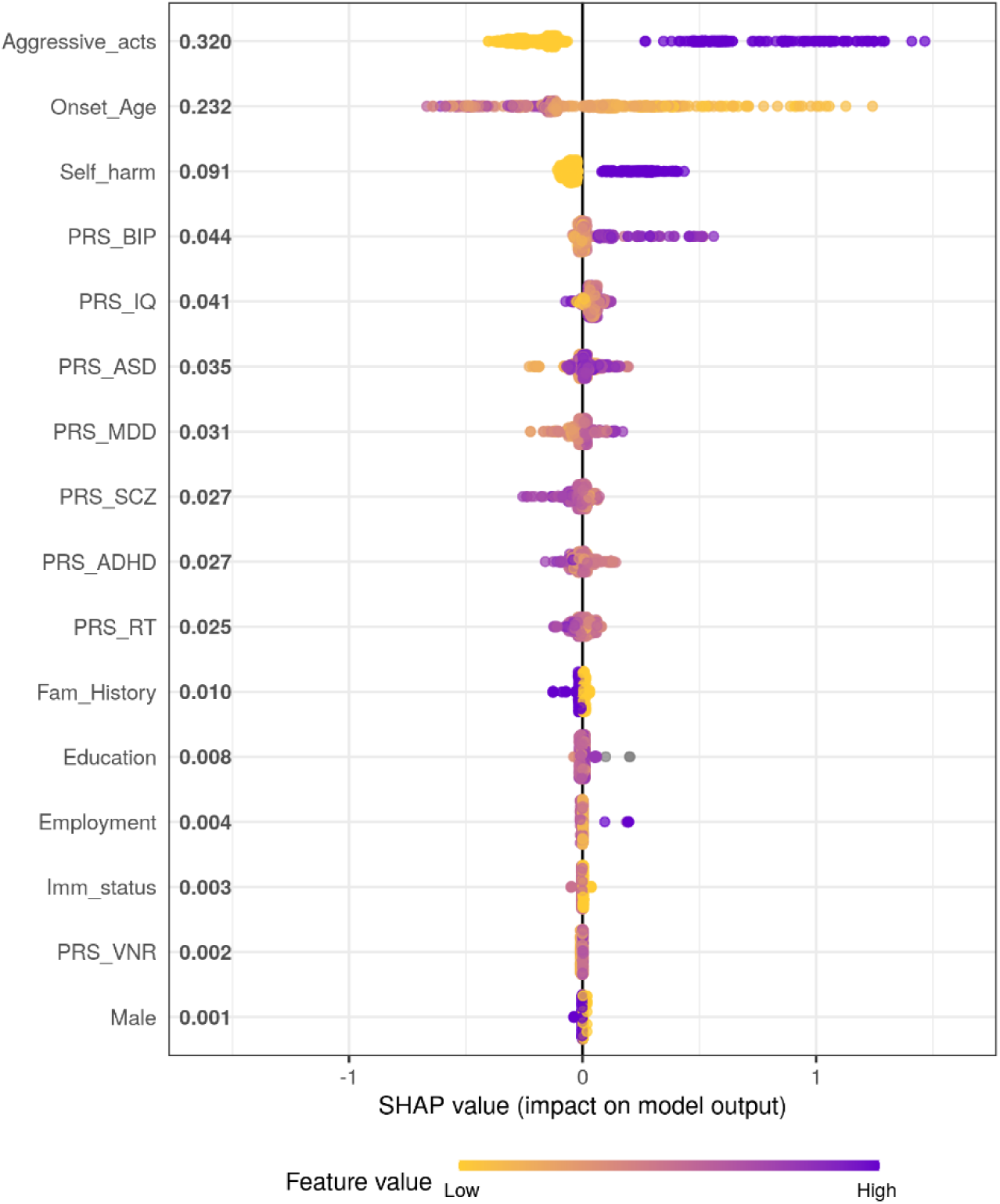
Mean Shapley value overall VarImp plot for prediction of total duration of hospitalization.

**Figure 15.**
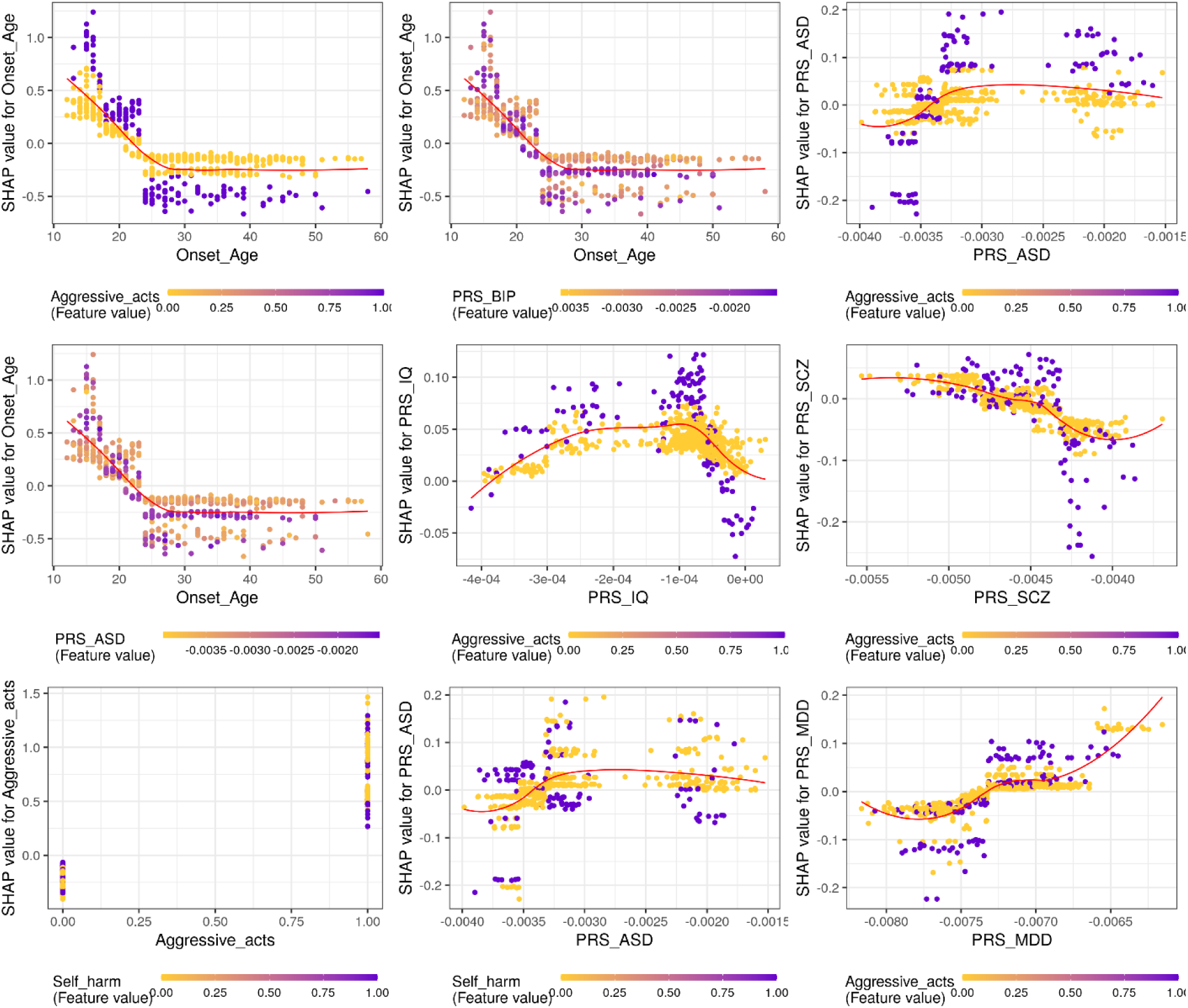
Shapley value interaction plot for the top 9 interaction pairs for prediction of total duration of hospitalization.

**Table 17.** Five-fold cross-validated prediction accuracy for prediction of total duration of hospitalization.

| **Fold** | **RMSE** | **R-Squared ($R^2$)** |
| --- | --- | --- |
| **1** | 4.561 | -0.035 |
| **2** | 3.342 | -0.115 |
| **3** | 2.864 | 0.111 |
| **4** | 2.527 | 0.081 |
| **5** | 2.020 | 0.098 |
| **Average** | 3.063 | 0.028 |

\*Remark: The negative $R^2$ is due to the use of adjusted $R^2$ .

**Table 18.**
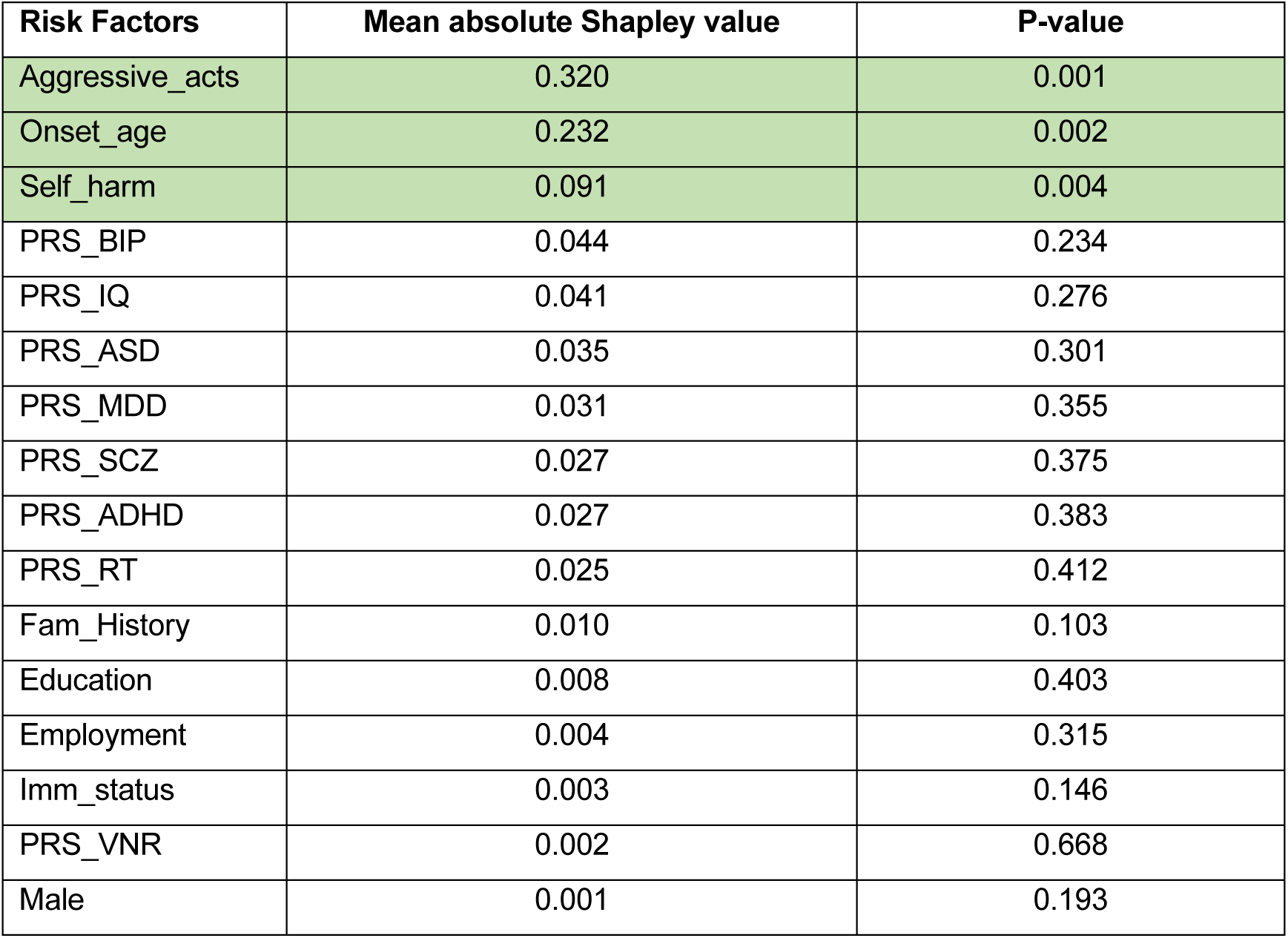
Permutation (N=500) P-value of mean absolute Shapley value for prediction of total duration of hospitalization.

## Discussion

To the best of our knowledge, this study is among the first GWAS examining clinical profiles in Chinese SCZ patients. Nevertheless, we made efforts to explore variants and genes across different ethnic groups. Conducting a meta-analysis that combines Chinese and European samples enhances statistical power and robustness of identified SNPs. The results of our studies are elaborated upon in the following sub-sections.

### Genome-wide association study on clinical phenotype and meta-analysis of GWAS results

The primary objective of this study was to identify genetic variants significantly associated with various clinical phenotypes in patients with SCZ across three cohorts, including both Hong Kong Chinese and European populations. In the Hong Kong cohorts, the GWAS analysis of the AAO phenotype revealed a significant association with a genetic variant rs142498233 on chromosome 7. This association was further validated as an independent genetic locus through clumping analysis. In the European cohort, the GWAS analysis of the AAO phenotype identified two significantly associated variants. After clumping analysis, three independent genetic loci on different chromosomes were identified. Meta-analysis across the three cohorts, which encompassing 1,500 SCZ patients, confirmed one significant signal (rs142498233) from the Hong Kong cohorts and two significant signals(rs185213255 and rs144645158) from the European cohort. Notably, Sada-Fuente *et al.* conducted a large GWAS analysis of AAO in a European cohort of 4,740 SCZ subjects, estimating the SNP-based heritability of AAO to be between 17% and 21% [24]. However, they did not identify any genome-wide significant loci in the European cohort. In contrast, our study integrated data from both Chinese and European cohorts, revealing several independent loci. This approach can be considered a cost-efficient method to identify significant associated genetic variants for clinical phenotypes in SCZ patients with moderate heritability. Additionally, our study identified, for the first time, a genetic variant rs60648049 on chromosome 9 that significantly associated with hospitalization count in cohort 1.

### Deciphering the genetic relationship of SCZ phenotypes with psychiatric disorders/traits

In this study, we introduce novel evidence supporting a shared genetic basis among four psychiatric disorders, four cognitive functions, and three SCZ clinical phenotypes. This shared genetic susceptibility is evidenced by the nominal genetic correlations observed between psychiatric traits and cognitive functions. Our study highlights the multifaceted nature of phenotypic manifestations, emphasizing the critical interplay between genetic predispositions, as indicated by PRS, and socio-demographic variables such as gender and educational level. The findings reveal a complex milieu in which both genetic makeup and socio-demographic attributes significantly influence the expression and severity of specific behaviours and conditions. In particular, our results highlight the considerable impact that educational attainment and gender differences have in modulating the effects of genetic predispositions on phenotypic expressions.

Additionally, the potential links between specific PRS (e.g., for IQ, prospective memory, ADHD) and phenotypes underscore the intricate genetic architecture underlying these manifestations. This complexity suggests the presence of a shared genetic basis that might either magnify or mitigate the effects of socio-demographic factors on these phenotypes, showcasing that genetic predisposition, through its complex interactions with environmental factors, plays a fundamental role in the manifestation and variability of these behaviours and conditions.

Other studies consistently reinforce the complex relationship between SCZ phenotypes and variables such as IQ [25], gender [26–28], and years of education [27, 29, 30], with aggressive behaviours being notably recognized as a significant phenotype within SCZ [31]. Our research postulates the existence of a shared genetic basis linking aggressive tendencies with reduced IQ among individuals diagnosed with SCZ. The study [25] conducted by Tilman Steinert and colleagues corroborate our findings, revealing a discernible reduction in IQ among SCZ patients exhibiting aggressive behaviours. This suggests that the progression of SCZ may lead to cerebral abnormalities that concurrently impair cognitive function and behavioural regulation, predisposing those with aggressive tendencies to a marked reduction in cognitive capabilities.

Our investigation further clarifies the gender-specific genetic correlations between aggressive behaviours and self-harm tendencies within the SCZ context. The prevalence of aggressive behaviours is reported more frequently among male SCZ patients than females, a finding consistently supported by numerous studies [26, 27]. This divergence is speculated to originate from variations in genetic expression and the effects of hormonal and physiological changes. Our analysis agrees with the conventional academic perspective [28], indicating an earlier onset of SCZ among males compared to females, which may significantly influence the assessment. The emphasis on gender-specific genetic differences is imperative for producing unbiased and accurate research outcomes. Given the variations in SCZ manifestation across genders—with males generally experiencing earlier onset and more severe aggressive behaviours—the integration of these distinctions into research methodologies is essential. Overlooking these crucial gender-based genetic variations could substantially alter the interpretation and applicability of findings, necessitating a sophisticated approach to SCZ research. This underscores the significance of incorporating gender as a key factor in exploring the genetic and developmental trajectories of SCZ, thereby fostering a more comprehensive and equitable understanding of the disorder.

These insights stress the paramount importance of meticulously considering gender-influenced genetic variations within SCZ research. We advocate for a deeper understanding and thorough examination of gender-associated disparities in disease presentation, thus propelling the field of genetic and developmental research related to SCZ forward.

Historically, the bulk of research into the characteristics and tendencies of SCZ has utilized survey questionnaires, a method that, while offering a general overview of disease traits, falls short in unravelling the genetic foundations of these characteristics or devising definitive, actionable interventions. Our study marks a significant advance in exploring the shared genetic basis between SCZ phenotypes in the Chinese cohorts and additional traits through genetic analysis.

This study employs generalized linear model (GLM) analysis to shed light on the nuanced interplay between genetic predispositions, as measured by PRS, and socio-demographic factors such as gender and educational level. This approach enhances our comprehension of the subtle influences on the severity and presentation of aggressive and self-harm behaviours, as well as the AAO in SCZ. Such insights provide a foundational basis for further investigations into targeted intervention strategies, contributing significantly to the discourse on the complex interrelations between genetics and socio-demographic factors in determining phenotypic outcomes.

### Genome-wide gene-based association analysis and transcriptomics-wide association study (TWAS) analysis

To further elucidate the biological significance of the associated variants, we conducted gene-based association analyses for clinical phenotypes. Although no gene was significantly associated with AAO, significantly enriched gene sets and pathways were identified using the nominally associated genes with AAO in GO sets and KEGG pathway enrichment analyses. Interestingly, the analyses revealed a significant association between the AAO phenotype and pathways including *alcoholic liver disease (hsa04936)*, *lipid and atherosclerosis (hsa05417)*, *non-alcoholic fatty liver disease (hsa04932)*, and *Diabetic cardiomyopathy (hsa05415)*.

Furthermore, by integrating genetically imputed gene expression data (whole brain tissue) and GWAS summary statistics, we identified several significantly associated genes with multiple SCZ-related phenotypes. These included the *DDIAS* gene in the frontal cortex region, the *GAS2* gene and *NBPF26* gene in the cerebellum region associated with the AAO phenotype, the *RIC8A* gene associated with aggressive acts in the putamen-basal-ganglia region, and the *ADGRE2* gene associated with hospitalization in the amygdala region. The *CARD14* gene encodes a caspase recruitment domain-containing protein and acts as a scaffolding protein that can activate the inflammatory transcription factor NF-kappa-B and p38/JNK MAP kinase signaling pathways [35]. The *ADGRE2* gene encodes a member of the class B seven-span transmembrane (TM7) subfamily of G-protein-coupled receptors, functioning as a cell surface receptor that binds to the chondroitin sulfate moiety of glycosaminoglycan chains and promotes cell attachment. In macrophages, the encoded protein regulates mast cell degranulation and facilitates the release of inflammatory cytokines, including IL-8 and TNF [36]. Moreover, the enrichment analysis of GO gene sets suggested that the molecular function of nuclease activity plays a role in the pathogenesis of the AAO phenotype. Genetic deficiencies, copy number variants, and single nucleotide variants of DNA repair nuclease-related genes have been linked to the 15q13.3 microdeletion/microduplication syndrome, which is associated with autism, schizophrenia, and epilepsy [37], further supporting the reliability of the enrichment analysis.

### Prediction of severity in schizophrenia patients

In this study, we developed ML models to predict the severity of clinical phenotype in patients with SCZ, using selected predictors and three different proxies as outcomes. The models achieved modest discrimination with a ROC-AUC of approximately 63.4%, comparable to findings from another study using a larger UKBB sample to discriminate SCZ cases and controls [38]. Predictive performance as measured by R^2^ and RMSE, was likewise modest but demostrated non-zero predictive value, thereby serving as a proof-of-concept for the application of the XGBoost model in predicting phenotype severity through the number and total duration of hospitalization.

Regarding the results for the mean Shapley value variable importance (VarImp) plot and permutation P-value of the mean absolute Shapley value, three factors were consistently identified as significantly worsening the SCZ phenotypes across different severity proxies: lower AAO, having aggressive behavior, and exhibiting self-harm acts. The consistency of these predictors suggests that they are robust indicators for predicting the clinical outcomes of SCZ patients. An earlier AAO has been found to be associated with increased symptom severity and poorer clinical outcome in SCZ. One study [3] revealed that this correlation originates from a shared genetic effect that influences both the developmental timing of SCZ onset and the severity of negative symptoms among SCZ individuals. Aggressive or violent behavior has been recognized as one of the clinical manifestations of SCZ, where an elevated risk of committing violent crimes observed in patients with SCZ spectrum disorders [4]. Additionally, patients exhibiting aggressive behaviors such as physical violence or threats are more likely to be diagnosed with a serious mental illness including SCZ [5]; Self-harm acts are strong indicators of distress and predictors of SCZ severity, where attempters of self-harm or suicidal acts have been reported to suffer from more severe SCZ symptoms than non-attempters [6].

Additionally, Shapley value analysis further identified the PRS score for BIP as an important predictor for SCZ severity. This can be explained by the substantial genetic overlap in shared risk alleles between SCZ and BIP. It has been suggested that higher PRS for BIP is associated with an increased risk and severity of SCZ [7]. The PRS for IQ was also found to increase the severity of SCZ, which was supported by another study [38] showing that the fluidic intelligence explained the highest variance in the fitted model for predict SCZ risk.

The interaction analysis plot also yielded some interesting results, indicating that the combinations of several predictors could affect the SCZ severity. Notably, the interaction term between AAO and aggressive acts exhibited a different pattern for severity prediction: An early AAO combined with the presence of aggressive acts elevated the severity of SCZ, as represented by all three proxies. In contrast, a late AAO combined with having aggressive acts alleviated this severity. This pattern can possibly be explained by the onset of SCZ later in life following a different clinical trajectory. Aggression in the context of late-onset SCZ may indicate a different underlying pathology or comorbid conditions when hospitalization is less frequently required. The integration of more sophisticated models may be further needed to enhance our understandings of how these predictors interact to influence SCZ severity.

These findings underscore the importance of specific clinical and genetic factors in predicting severity phenotypes in subjects from the Chinese SCZ cohorts, highlighting the potential for early intervention strategies targeting these risk factors to mitigate severe outcomes. In conclusion, our study provides valuable insights into the predictors of SCZ severity and demonstrates the efficacy of mML approaches, especially XGBoost and Shapley value analysis, in clinical prediction tasks. These tools offer a promising avenue and form a basis for further studies to improve patient outcomes through targeted interventions.

## Limitations

### Genome-wide association study on clinical phenotype and meta-analysis of GWAS results

The present study has several limitations that should be acknowledged. Firstly, the sample size of each individual cohort of SCZ patients was relatively small, ranging from 387 to 702 subjects. This limitation may weaken the reliability of the GWAS results. To address this issue, the present study employed a meta-analysis approach to integrate the GWAS summary statistics of the three cohorts, thereby improving the statistical power. Furthermore, by combining genetic information from both Chinese and European populations, this study has identified several novel significant genetic variants. However, it is important to note that the inconsistent collection methods for clinical phenotypes in different cohorts hindered the ability to perform meta-analysis for certain phenotypes, including hospitalization and SCZ episodic. Furthermore, all genome-wide significant SNPs should be considered putative, requiring replication in larger independent AAO GWAS.

### Deciphering the genetic relationship of SCZ phenotypes with psychiatric disorders/traits

It is crucial to acknowledge the limitations imposed by our relatively small sample, which may introduce biases into the results. In addition, the PRS were derived from European GWAS and applied to our Chinese cohort, a mismatch of ancestry that may reduce the statistical power and thus affect the accuracy of the regression models. Lastly, including all PRS simultaneously in a linear regression model may raise concerns about multicollinearity, as some PRS are genetically correlated. In summary, it is essential to adopt and implement a broader spectrum of methods capable of more accurately deciphering the genetic signals associated with SCZ.

### Genome-wide gene-based association analysis and transcriptomics-wide association study (TWAS) analysis

The relatively small number of associated genes obtained from the tissue-specific imputed gene expression association analyses may have resulted in a lack of significantly enriched pathways after multiple correction testing.

### Prediction of severity in schizophrenia patients

However, as the relatively small sample size may introduce bias to the fitted prediction model, it still needs to be validated in more diverse and large study samples before clinical applications. Future work should also aim to refine these predictive models by incorporating additional clinical and genetic data. Furthermore, to enhance our understanding of how the predictors interact to influence SCZ severity, more sophisticated models may be needed.

## Conclusions

This study represents one of the first batch GWAS examining clinical profiles in Chinese schizophrenia (SCZ) patients and highlights the benefits of incorporating data from multiple ethnic groups. By combining Chinese and European samples, we increased statistical power and identified robust candidate SNPs associated across various SCZ phenotypes.

We also provided evidence of a shared genetic basis among psychiatric disorders, cognitive functions, and SCZ phenotypes, demonstrating suggestive trends toward genetic correlations between psychiatric traits and cognitive functions. This study emphasizes the influence of socio-demographic variables, such as gender and educational level, on the expression and severity of SCZ phenotypes. The exploratory findings underscore the complex interplay between genetic predispositions and environmental factors, influencing the manifestation and variability of SCZ-related behaviors and conditions.

Whole-genome gene-based analyses implicated genes like *WSB2*, *DDIAS*, and *ADGRE2* in AAO, aggressive behavior, and hospitalization, respectively. Transcriptome-wide analysis highlighted the role of telomeric regions and nuclease activity in the pathogenesis of AAO.

Moreover, we developed ML models to predict the severity of SCZ phenotypes, achieving moderate discrimination and acceptable predictive performance. Factors such as lower AAO, aggressive behavior, and self-harm acts were identified as robust predictors of SCZ severity. The study also highlighted the importance of PRS for bipolar disorder and IQ in predicting SCZ severity.

In conclusion, our findings provide novel insights into the genetic architecture of SCZ phenotypes and their relationship with other psychiatric traits. The identification of predictive factors paves the way for targeted interventions aimed at mitigating severe outcomes. This multi-pronged approach combining genomics, transcriptomics, and ML holds promise for unraveling the complex etiology of SCZ and improving patient management strategies.

## Data Availability

The data that support the findings of this study are available upon request from the corresponding authors, Hon-Cheong SO and Simon Sai-Yu LUI. The data is not publicly available because it contains information that could compromise the privacy of research participants.

## Acknowledgements

This study was supported by the Health and Health Services Research Fund from the Food and Health Bureau, Hong Kong Special Administration Region. We want to thank the Health and Medical Research Fund (Project no. **07180376**), HKSAR, for the support of this project. We are also grateful to all the staff of the EASY team in Castle Peak Hospital (CPH), HKU Dept of Psychiatry, and the participants of this study, for their valuable contributions.

## Supplementary Tables

[Download link](https://drive.google.com/file/d/12RmA-bigfcWxlbMXxNs96XGgl1PEw_EJ/view?usp=sharing) to the Supplementary Tables ST1 to ST10.

## Notes

### Competing Interest Statement

The authors have declared no competing interest.

### Funding Statement

Health and Medical Research Fund (Project no. 07180376), Hong Kong SAR.

### Author Declarations

Ethics committee/IRB of the Joint Chinese University of Hong Kong - New Territories East Cluster (CUHK-NTEC) Clinical Research Ethics Committee or the New Territories West Cluster Research Ethics Committee (NTW CREC) gave ethical approval for this work.

